# A target trial emulation of adherence to Canada’s Food Guide 2019 recommendations in older adults

**DOI:** 10.1101/2025.07.03.25330841

**Authors:** Didier Brassard, Nancy Presse, Stéphanie Chevalier

## Abstract

The 2019 Canada’s Food Guide (CFG) may not be tailored for older adults, since it provides universal recommendations. In community-dwelling adults aged 67-84 years and compared with habitual diet and physical activity, our objective was to estimate the 3-year difference in muscle strength, physical function, cardiometabolic health and cognitive health score according to adherence to CFG recommendations (CFG), enhancements with additional protein foods (CFG-PRO), physical activity (PA) and both (CFG-PLUS). Longitudinal non-experimental data from the NuAge study (2003-2008 in Québec, Canada) were used to emulate a 3-year target trial. Data was collected at annual in-person follow-up visits. The hypothetical interventions were modelled using the parametric g-formula assuming no unmeasured confounding, no measurement error and correct models. 1561 participants were eligible. Compared with no intervention, adherence to the CFG intervention would have increased quadriceps strength by 0.8 kg (95%CI: 0.0, 1.7), walking speed by 0.03 m/s (95%CI: 0.00, 0.05) and reduced waist circumference by 1.0 cm (95%CI: -1.7, -0.3). Estimates were similar for the CFG-PRO intervention. Compared with CFG alone, the CFG-PLUS intervention improved to a greater extent walking speed (vs. CFG, +0.06 m/s; 95%CI: 0.03, 0.08) and waist circumference (vs. CFG, -0.8 cm; 95%CI: -1.5, -0.2). Under strong assumptions, compared with no intervention leading to declines in most outcomes, adherence to CFG recommendations over 3 years would have attenuated declines in strength and walking speed in older adults. Greater benefits were achieved when protein food intake and physical activity were increased in addition to CFG adherence.

## Introduction

Canada’s Food Guide (CFG) was revised in 2019 to “*promote healthy eating and overall nutritional well-being*” based on the latest evidence (1). More precisely, the underpinning literature for CFG focuses on chronic disease reduction such as cancer, cardiovascular disease, and type 2 diabetes (2). The recommendations are mostly qualitative (e.g., “eat more of …”), focus on plant-based food sources (i.e., vegetables and fruits, whole-grain foods, plant-based protein foods, unsaturated fats and oils) and are depicted using a plate (1). CFG recommendations are universal to all individuals aged 2 years and older (1).

In Canada, the proportion of adults aged 65 years and older has been increasing rapidly to 19.5% in 2025 (3). Older adults are more vulnerable to inadequate dietary intakes due to social and physiological changes (4). For example, one third of community-dwelling older adults are at nutritional risk (5, 6). In turn, older adults affected by malnutrition have an increased risk of frailty (7) and sarcopenia (8), which affects their autonomy and increases their risk of institutionalization (9). Evidence to date supports the adequacy of CFG recommendations for cardiovascular disease prevention (10). Yet, their appropriateness for fulfilling nutritional requirements and contributing to muscular and cognitive health, which are key health outcomes among older adults to maintain autonomy (8, 11), has not been documented comprehensively. Data from the 2015 Canadian Community Health Survey (CCHS) – Nutrition showed that high adherence to CFG recommendations on healthy food choices was insufficient to mitigate calcium and folate inadequacy, and insufficient to support optimal protein intakes, in older adults (12). A growing consensus also supports higher than recommended protein intakes for older adults, such as a recommended daily intake of 1.0 to 1.2 g/kg (13). Adherence to CFG recommendations may be insufficient to mitigate the substantial inadequacy to those higher protein requirements among older adults from Canada (12). For example, more than 18% and 50% of Canadians aged 65 y or older would have protein intakes below the current recommended daily allowance of 0.8 g/kg and the higher recommended intake of 1.0 g/kg, respectively, (12). Thus, the universal CFG recommendations may not be optimal for older adults. Finally, only 33% of older adults in Canada meet moderate-to-vigorous physical activity recommendations (14). Given the benefits of physical activity and protein intake to improve muscle function and strength (13, 15), how the adherence to CFG recommendations alone or combined with physical activity contributes to favorable health outcomes should be examined.

A large randomized controlled trial (RCT) would ideally document the effect of adhering to CFG recommendations. However, nutritional intervention studies are particularly challenging to conduct due to blinding and long-term adherence issues (16, 17). Indeed, strictly controlling dietary intakes over a long period is challenging in free-living conditions. As a first step, using observational data is an appropriate alternative. Using the target trial framework (18, 19), our objective was to estimate the difference in key health outcomes among eligible community-dwelling older adults from the NuAge study, at the end of follow-up, had they followed the intervention vs., instead, had they maintained their diet and physical activity habits. More precisely, we aimed to estimate the end of follow-up difference in muscle strength, physical function as well as cardiometabolic markers and cognition following these hypothetical interventions over 3 years, compared with no intervention: 1) the effect of adhering to CFG 2019 recommendations (CFG); 2) same as 1, with additional protein foods (CFG-PRO); 3) increasing physical activity without diet modifications (PA); 4) same as 1, with additional protein foods and increasing physical activity (CFG-PLUS). Of note, “no intervention” corresponds to the habitual diet and physical activity habits as reported by participants. We hypothesized that adherence to CFG recommendations would improve cardiometabolic and cognitive health but that only the enhanced recommendations, i.e., those including additional protein foods and physical activity, would improve muscle strength and function compared with no intervention.

## Methods

### Design and participants

Complete details about the present study protocol have been published elsewhere (19), including the justification for the hypothetical diet and physical activity intervention, the hypothesized relationships between variables and the thorough description of the target trial. **Figure S1** presents an overview of the study design. Briefly, participants were adults aged 67-84 y from the longitudinal NuAge Database and Biobank (20, 21). The NuAge participants were identified in 2003 based on a random sample of 36,183 males and females from the Quebec health insurance database, stratified for age and sex (20). A total of 1793 participants were recruited from three regions in the Province of Quebec, Canada (Sherbrooke, Montreal, or Laval) between 2003 and 2005 at baseline and followed over three years. Data collection was completed in June 2008. All participants of the NuAge study provided informed consents. A subset of 1753 participants (98%) further agreed to have their data included in the NuAge Database and Biobank used in the present study (20).

We aimed to explicitly emulate the protocol of a target trial (18, 22) using non-experimental data from the NuAge study (Table 1).

**Table 1:**
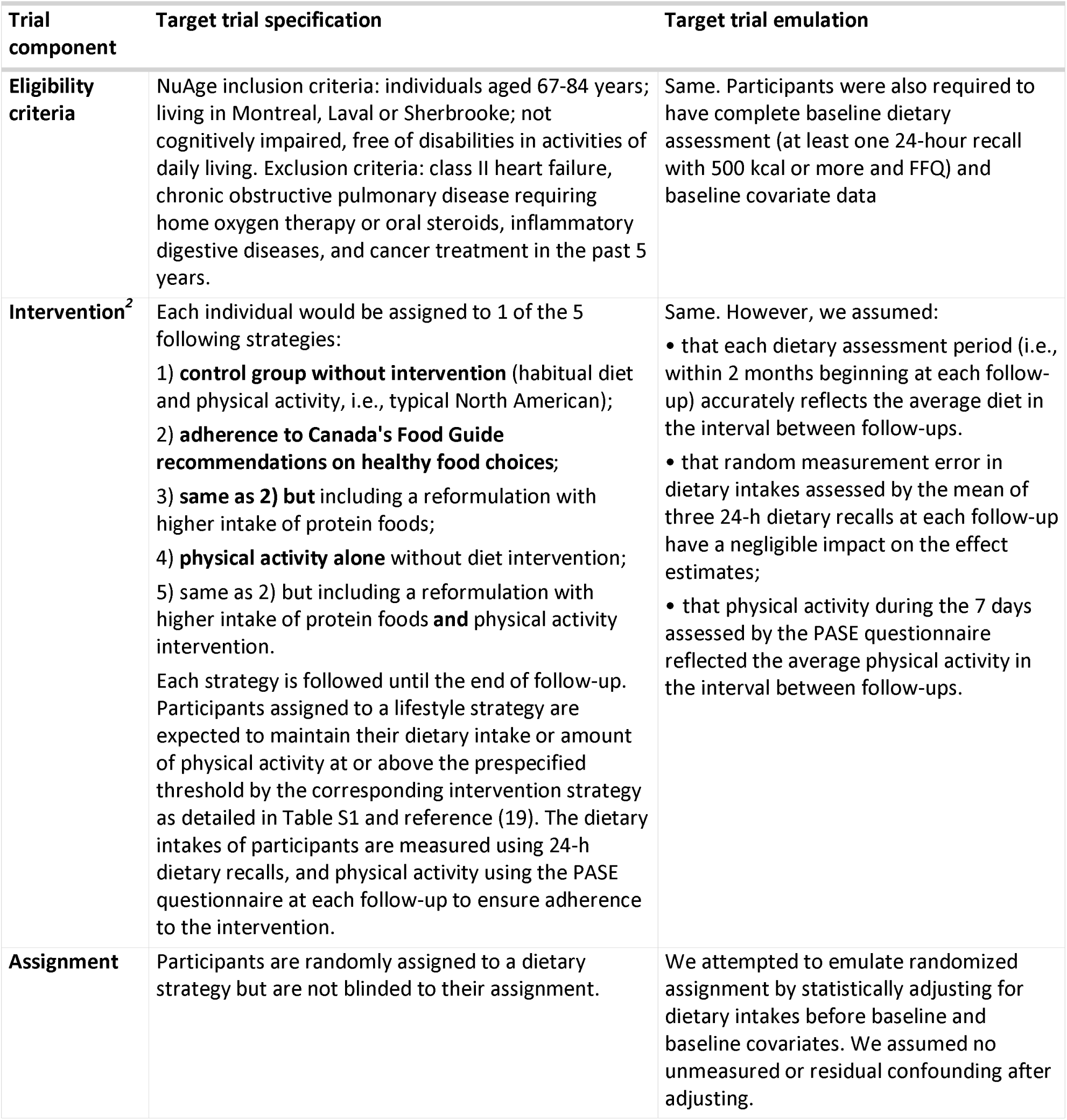
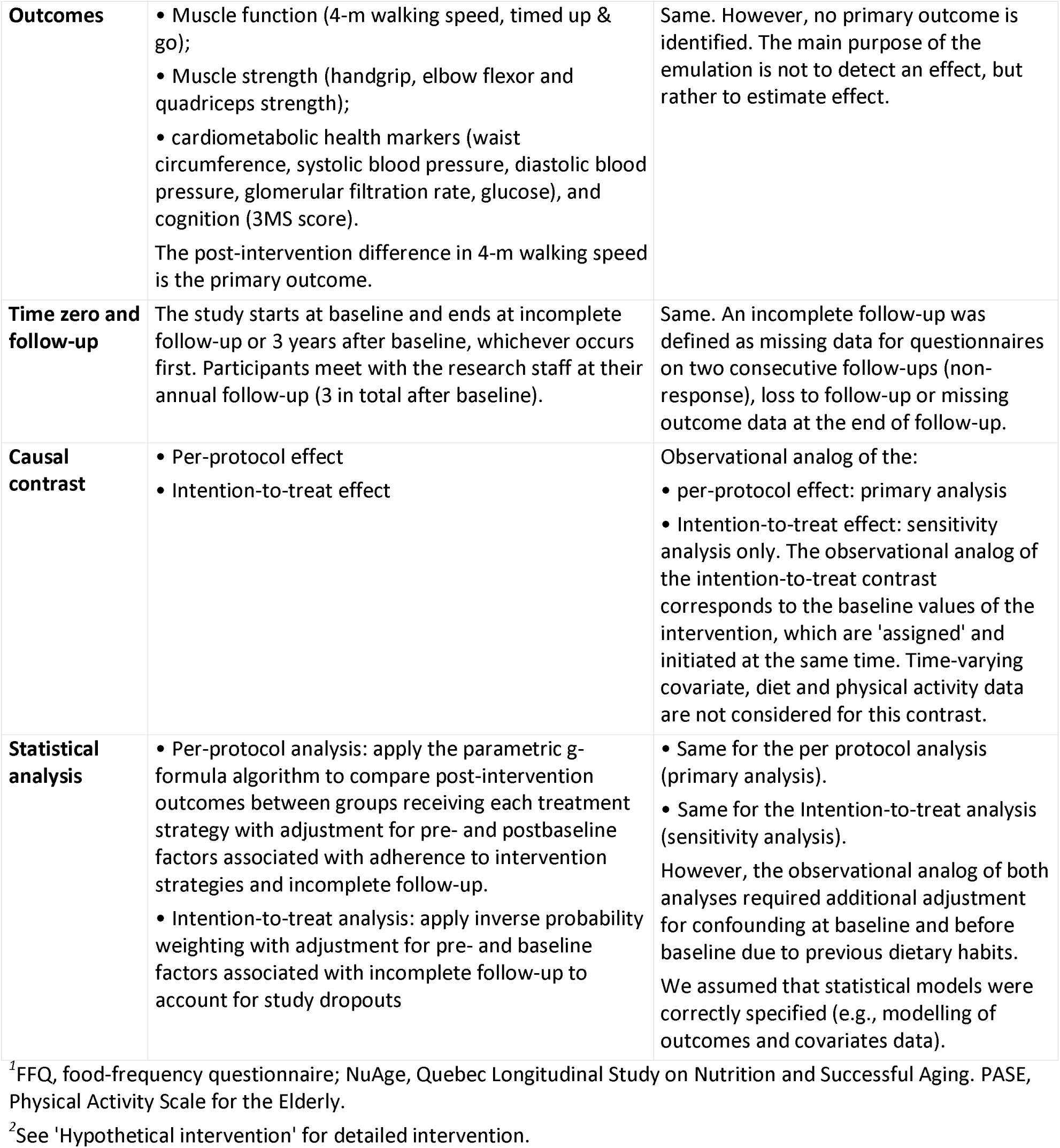
Emulation of a dietary intervention target trial using non-experimental data from the NuAge study.

### Exposure: Diet and physical activity

#### 24-hour dietary recalls (main exposure)

Dietary intakes at baseline and at each annual follow-up (up to three) were assessed using interviewer-administered 24-h dietary recalls (21). Up to three non-consecutive 24-h dietary recalls were collected at each time, including one face-to-face interview during the annual assessment and two unannounced interviews by telephone within two months. The interviews were administered according to the United States Department of Agriculture multiple-pass method (23). Portion size estimation was facilitated using portion-size models and pictures of standardized food portions. For the present study, reported food and beverages were classified according to the food categories corresponding to each component of the hypothetical diet intervention (complete detail in Supplemental Methods – hypothetical intervention). The categories were vegetables and fruits, whole-grain foods, plant-based protein foods, animal-based protein foods, unsweetened milk and plant-based beverages with protein, unsaturated fats and oils, water and other healthy beverages. Foods and beverages not recommended were also classified including other low nutritive value foods, sugary drinks and alcohol, and non-whole grain foods (19). The number of reference amount (RA) was calculated for each dietary item. Similar to a standardized portion size, RA corresponds to the amount of food in grams typically consumed at one sitting in Canada (24). Mean intakes in RA were then calculated at each follow-up for each category and participant based on all 24-h dietary recalls completed with 500 kcal or more (19).

#### Food frequency questionnaire (pre-baseline dietary intakes)

Dietary intake in the year prior to the baseline were assessed using a semi-quantitative food frequency questionnaire (FFQ) (25). Portion size estimation was facilitated using pictures of sample servings throughout the questionnaire. To have standardize portion size similar to the 24-h dietary recall data, intake data for each question were converted to Reference Amount (RA) per day. Then, the sum of intakes was calculated among the food and beverage categories of the hypothetical intervention. FFQ with reported energy intakes <800 kcal and >4200 kcal (males) or <500 and >3500 kcal (females) were excluded as well as FFQ with >25% missing responses.

#### Physical activity

Physical activity over the past 7 days was estimated using the Physical Activity Scale for the Elderly (PASE) questionnaire (26, 27). The questionnaire was completed at the same assessment visit as the first 24-h dietary recall at each yearly follow-up. The amount of light intensity or higher physical activity in minutes per day was derived based on individual PASE questions.

#### Hypothetical intervention

**Table S1** presents the detailed dietary intervention including the pre-specified thresholds for each dietary component and physical activity. For the hypothetical intervention, participants would be expected to meet or exceed each target dietary intakes for each dietary component as well as the target amount of physical activity for intervention involving physical activity (19). To avoid increasing the amount of food consumed, the dietary intervention would be implemented as a “substitution” (19). For example, to meet the target vegetables and fruits intake, a male participant could be instructed to increase their intake from 3 servings (as reported using 24-h dietary recalls) to 6 servings (target in Table S1), while proportionately decreasing their intakes of low nutritive value foods, sugary drinks and alcohol, saturated fats and oils, or non-whole grains. This hypothetical substitution intervention is implemented in the analysis by adjusting for total food intakes at each follow-up, which included total food and total beverage intakes (19, 28–30). Total food intake was included as a time-varying covariate in the intervention modelling (Statistical Analyses).

### Covariates

Sociodemographic, lifestyle, health and anthropometric data were collected using interviewer-administered standardized questionnaires and procedures at baseline and at each follow-up (20, 21). Potential data-entry errors were mitigated using predefined fields and outlier flags. Trained registered dietitians and nurses conducted the interviews (21).

Covariates were selected based on background knowledge and causal directed acyclic graph to account for confounding due to prior dietary habits (before baseline), individual factors, the propensity towards health-seeking behaviors and disease burden (19). The pre-baseline covariates were intakes (RA/day) of vegetables and fruits, whole grains, refined grains, animal-based protein foods, plant-based protein foods, milk, sugary drinks and alcohol, other low nutritive value foods, oils and fats that are saturated or unsaturated. The baseline covariates were age (years), sex (male or female), region (Sherbrooke, Montreal, or Laval), education (years), height (m), and former cancer (yes/no). The time-varying covariates were intakes of unsweetened drinks (RA/day), total dietary intakes (RA/day), measured bodyweight (kg), light to high intensity physical activity (minutes/day; in diet-only interventions), living alone (yes/no), the number of medication, the use of vitamins, minerals or natural health product (yes/no), the consumption of alcohol in the prior month (yes/no), current smoking (yes/no), the presence of chronic disease (diabetes, high blood pressure or heart disease) and being “excused” from adhering to the intervention (yes/no; see below for details). Data on time-varying covariates collected at each annual follow-up were used in the analysis, as described below.

### Outcomes

Key outcomes related to muscle strength, muscle function as well as cardiometabolic and cognitive health were selected based on availability in the NuAge Database and Biobank and relevance for older adults (19–21). All outcomes were measured using standardized protocols (21) (**Table S2)**. The muscle strength outcomes were handgrip, elbow flexor and quadriceps strength. The muscle function outcomes were walking speed and timed “Up & Go”; the cardiometabolic health outcomes were waist circumference, blood pressure (systolic, diastolic), blood glucose, estimated glomerular filtration rate (eGFR); and cognition was assessed using the modified Mini-Mental State Examination (3MS) score. Blood samples were obtained in the morning after an overnight fast. Participants were instructed not to eat or drink (except reasonable amount of water), smoke, or chew gum after 19:00 the evening before. The end of follow-up values were used as outcomes.

### Statistical analyses

All analyses were conducted in R version 4.4.3 (2025-02-28; R foundation) and the gfoRmula package v1.1.1 (31).

#### G-formula algorithm

The observational analog of the per protocol contrast for the hypothetical interventions, the primary analysis, was estimated using the parametric g-formula algorithm (31, 32). Briefly, the g-formula algorithm uses parametric models and Monte Carlo simulations to estimate causal effects in longitudinal data including time-varying exposures and confounders (Supplemental Methods – g formula algorithm). In the present study, the g-formula was implemented to estimate the effect of adhering to the hypothetical intervention pre-specified in Table 1 and Table S1, while accounting for confounding and loss to follow-up. **Table S3** describes the modelling approach for each variable.

Variance was estimated using 500 bootstrap samples (percentile method). The mean difference between outcomes under the hypothetical intervention and “no intervention” (natural course) estimated the effect of the intervention. To assess the added value of consuming more protein foods and including physical activity, the between-intervention differences were also estimated for CFG-PLUS compared with CFG alone.

The hypothetical lifestyle intervention could be difficult or impossible to implement for participants with a high disease burden, especially for the physical activity component (19, 33). To mitigate confounding by disease burden, participants who received a cancer diagnosis, were hospitalized (>1 day) or were instructed to bed rest (>1 day) during the year between follow-ups were “excused” from adhering to the intervention. In other words, “excused” individuals were considered in the intervention group data to estimate outcome values, while leaving their dietary intakes or amount of physical activity unchanged.

#### Missing time-varying data

The previous value of missing time-varying data at a given follow-up was carried forward once. Participants were considered as having incomplete follow-up if questionnaire non-response occurred at 2 consecutive follow-ups. **Table S4** presents the proportion of missing time-varying diet, physical activity and covariates across follow-ups in eligible NuAge participants.

#### Subgroup and Sensitivity analyses

Stratified analyses by sex were conducted to assess the impact of biological sex and, to some extent, gender.

Sensitivity analyses were conducted to challenge key assumptions of the causal effect estimation process. First, the effect of the hypothetical interventions on albumin concentrations was estimated as a negative control outcome (34). Albumin was selected as a marker of overall health that should not be strongly modified by the hypothetical intervention, but that may otherwise reflect residual or unmeasured confounding by disease severity (35). Second, the impact of within-individual random errors associated with 24-h dietary recalls was assessed by comparing the uncorrected effect estimates with corrected effect estimates using the National Cancer Institute multivariate method (19, 36) (Supplemental Methods – Sensitivity analysis to assess the impact of measurement error). Third, to assess model misspecification in the g-formula algorithm, predicted values of time-varying variables under no intervention, as estimated by the g-formula, were compared to those estimated by inverse probability weighting (37).

## Results

### Study participants

Among 1753 total NuAge participants, 1561 were eligible at baseline (**Figure 1**). The characteristics of eligible and excluded NuAge participants (n=192) are shown in **Table S5. Table 2** presents baseline characteristics of eligible participants across adherence score to CFG. Among eligible participants, 276 (17.7%) were lost during follow-up.

**Figure 1:**
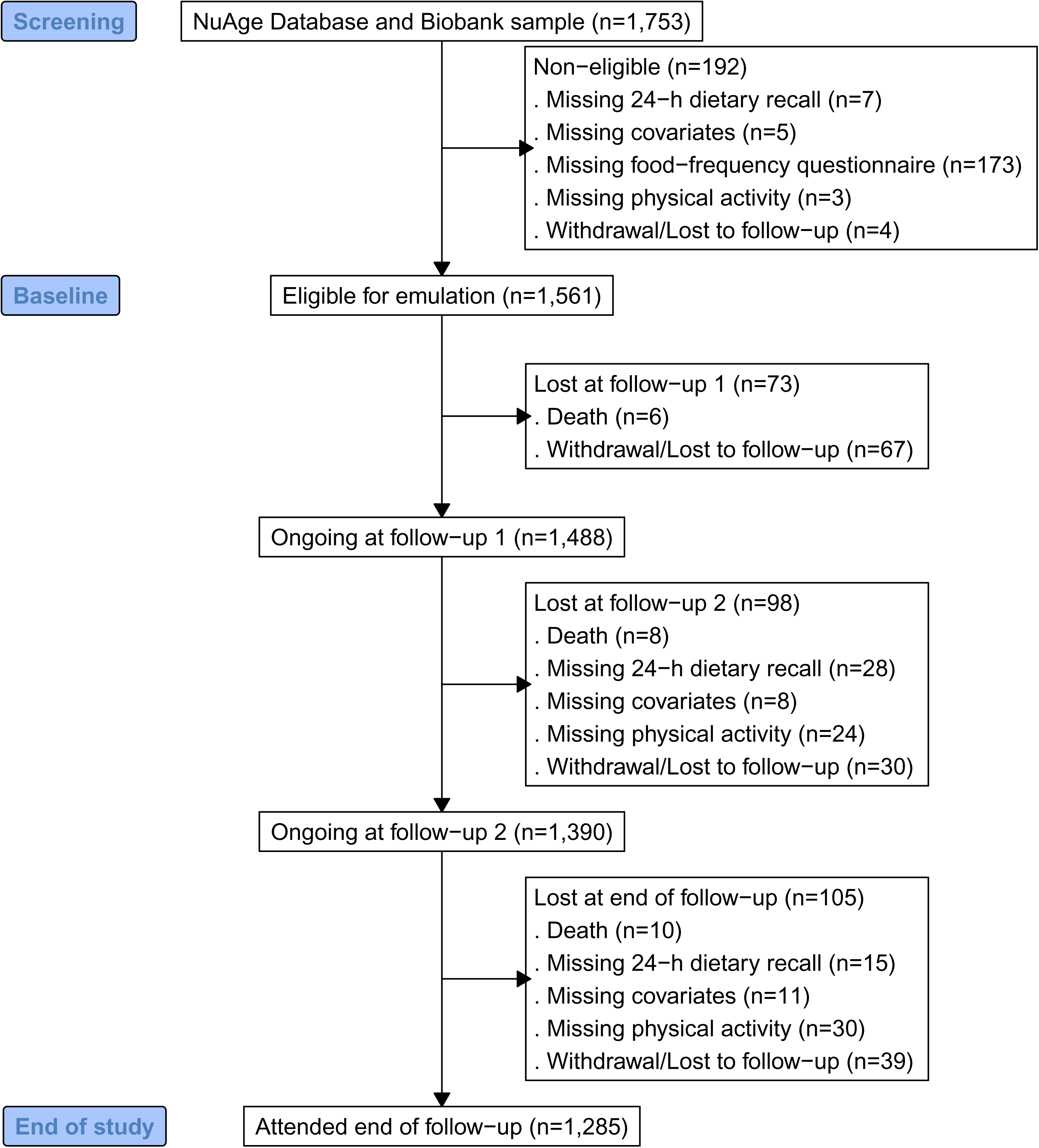
Flowchart of participants

**Table 2:** Baseline characteristics of eligible NuAge participants across quarters of HEFI-2019 total score ^1^.

| Characteristic | Overall, N = 1,561 | Quarters of total HEFI-2019 score |  |  |  |
| --- | --- | --- | --- | --- | --- |
|  |  | Q1 (min to 40.2), N = 384 | Q2 (>40.2 to 46.9), N = 388 | Q3 (>46.9 to 53.4), N = 394 | Q4 (>53.4 to max), N = 395 |
| Age, years | 74 (4) | 75 (4) | 74 (4) | 74 (4) | 74 (4) |
| Females | 809 (52%) | 161 (42%) | 182 (47%) | 219 (56%) | 247 (63%) |
| **Region** |  |  |  |  |  |
| Sherbrooke | 799 (51%) | 222 (58%) | 206 (53%) | 195 (49%) | 176 (45%) |
| Montréal | 448 (29%) | 84 (22%) | 95 (24%) | 115 (29%) | 154 (39%) |
| Laval | 314 (20%) | 78 (20%) | 87 (22%) | 84 (21%) | 65 (16%) |
| Education, years | 12 (4) | 12 (4) | 12 (5) | 11 (4) | 12 (4) |
| Household income, \$CAD<sup>2</sup> | 33,100 [26,900] | 31,700 [24,725] | 33,400 [27,450] | 35,000 [25,300] | 30,950 [29,375] |
| (Missing) | 201 | 52 | 57 | 47 | 45 |
| **Marital status** |  |  |  |  |  |
| Single | 167 (11%) | 43 (11%) | 50 (13%) | 37 (9.4%) | 37 (9.4%) |
| Widowed | 323 (21%) | 80 (21%) | 61 (16%) | 83 (21%) | 99 (25%) |
| Married | 944 (60%) | 232 (60%) | 245 (63%) | 246 (62%) | 221 (56%) |
| Separated, divorced | 127 (8.1%) | 29 (7.6%) | 32 (8.2%) | 28 (7.1%) | 38 (9.6%) |
| Living alone | 494 (32%) | 108 (28%) | 117 (30%) | 119 (30%) | 150 (38%) |
| **Ethnicity** |  |  |  |  |  |
| Caucasian | 1,540 (99%) | 381 (99%) | 385 (99%) | 385 (98%) | 389 (98%) |
| Non-Caucasian | 21 (1.3%) | 3 (0.8%) | 3 (0.8%) | 9 (2.3%) | 6 (1.5%) |
| Body mass index, kg/m<sup>2</sup> | 27.8 (4.4) | 27.9 (4.4) | 28.2 (4.6) | 28.0 (4.5) | 27.0 (4.2) |
| **Body mass index category** |  |  |  |  |  |
| Healthy | 416 (27%) | 97 (25%) | 91 (23%) | 103 (26%) | 125 (32%) |
| Overweight | 743 (48%) | 179 (47%) | 183 (47%) | 187 (47%) | 194 (49%) |
| Obesity | 402 (26%) | 108 (28%) | 114 (29%) | 104 (26%) | 76 (19%) |
| Total light or higher intensity activities, minutes/day<sup>2</sup> | 13 [39] | 6 [32] | 15 [39] | 10 [39] | 19 [39] |
| Meeting physical activity guidelines (150 minutes/week) | 404 (26%) | 70 (18%) | 98 (25%) | 111 (28%) | 125 (32%) |
| **Smoking status** |  |  |  |  |  |
| No | 809 (52%) | 178 (46%) | 192 (49%) | 220 (56%) | 219 (55%) |
| Formerly | 647 (41%) | 168 (44%) | 171 (44%) | 156 (40%) | 152 (38%) |
| Yes | 105 (6.7%) | 38 (9.9%) | 25 (6.4%) | 18 (4.6%) | 24 (6.1%) |
| Alcohol consumption in the last month | 1,265 (81%) | 308 (80%) | 334 (86%) | 324 (82%) | 299 (76%) |
| Vitamin, mineral or natural health product consumption | 492 (32%) | 94 (24%) | 117 (30%) | 136 (35%) | 145 (37%) |
| Difficulty chewing or meal interruption due to mouth or teeth problems | 301 (19%) | 82 (21%) | 70 (18%) | 81 (21%) | 68 (17%) |
| Self-reported high blood pressure | 733 (47%) | 196 (51%) | 172 (44%) | 182 (46%) | 183 (46%) |
| Self-reported diabetes | 165 (11%) | 29 (7.6%) | 46 (12%) | 41 (10%) | 49 (12%) |
| Self-reported heart problems | 342 (22%) | 98 (26%) | 77 (20%) | 83 (21%) | 84 (21%) |
| Number of medications<sup>2</sup> | 4 [5] | 4 [5] | 4 [5] | 4 [4] | 4 [5] |
| Past or current hormone replacement therapy (females only) | 451 (56%) | 75 (47%) | 105 (58%) | 132 (60%) | 139 (56%) |
| **Dietary intakes prior to baseline, RA/d<sup>3</sup>** |  |  |  |  |  |
| Vegetables and fruits | 5.3 (2.6) | 4.5 (2.3) | 4.9 (2.5) | 5.5 (2.4) | 6.2 (3.0) |
| Whole-grain foods | 1.4 (0.9) | 1.0 (0.9) | 1.4 (0.9) | 1.5 (0.9) | 1.7 (0.9) |
| Non-whole-grain foods | 1.5 (1.1) | 1.9 (1.2) | 1.6 (1.1) | 1.5 (1.0) | 1.1 (0.9) |
| Animal-based protein foods | 2.1 (1.0) | 2.1 (1.0) | 2.1 (1.0) | 2.1 (0.9) | 2.2 (1.0) |
| Plant-based protein foods | 0.7 (0.7) | 0.5 (0.6) | 0.6 (0.5) | 0.7 (0.7) | 0.9 (0.8) |
| Low nutritive value foods | 2.0 (1.6) | 2.9 (1.8) | 2.2 (1.5) | 1.8 (1.4) | 1.3 (1.1) |

<sup>1</sup>Values are mean (SD) unless otherwise indicated and frequency (column percentage). The HEFI-2019 total score reflects overall adherence to Canada's Food Guide recommendations on healthy food choices (range 0-80). HEFI-2019 quartiles were based on the raw distribution of the average of intake data from all 24-h dietary recalls completed. The score was used solely to describe the sample and not for the diet intervention modeling. HEFI-2019, Healthy Eating Food Index 2019; IQR, interquartile range; max, maximum value; min, minimum value; Q, quarter; RA, reference amount (amount of food typically consumed at one sitting in Canada (24)).
<sup>2</sup>Median [IQR]
<sup>3</sup>Dietary intakes were measured with a food-frequency questionnaire at baseline (see Methods) and reflect usual intakes in the year prior to baseline.

Median (Q1, Q3) follow-up time was 3.0 years (2.9, 3.1). Compared with participants below the first quarter of adherence score to CFG, participants in the highest quarter included more females, had a lower body mass index, were more likely to report doing 150 minutes of physical activity per week, to report not smoking, to consume vitamin or mineral supplements, or natural health products.

**Tables S6, S7 and S8** show respectively dietary intakes across follow-ups, the proportion of participants meeting intervention targets and dietary intakes after the hypothetical intervention. **Table S9** presents the observed values of outcomes and missing data across follow-ups in eligible NuAge participants.

### Effect of hypothetical interventions

The mean proportions of participants who had their reported dietary intakes or amount of physical activity modified across follow-ups were 91% for the CFG intervention, 92% for the CFG-PRO intervention, 55% for the PA intervention and 92% for the CFG-PLUS, respectively. The observed proportion of participants which were excused from adhering to the hypothetical interventions was 12% across follow-ups.

**Figures 2-4** show the difference in muscle strength, function and cardiometabolic health outcomes, respectively, after the hypothetical interventions at the end of follow-up.

**Figure 2:**
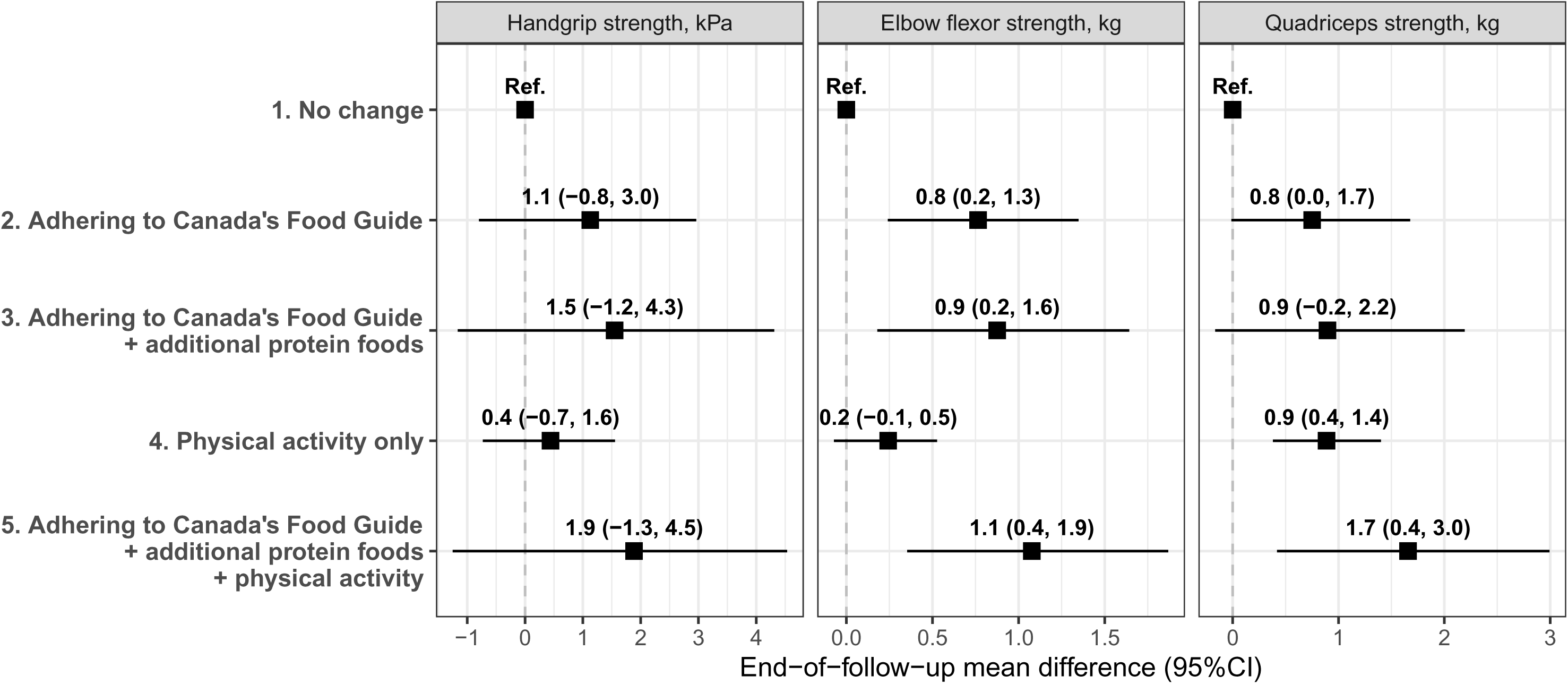
Mean difference in muscle strength outcomes after the 3-year hypothetical interventions, compared with no hypothetical intervention, in eligible NuAge participants. Effect estimates were obtained using the g-formula algorithm (see Methods). Physical activity corresponds to that of light intensity or higher. Models were adjusted for the covariates age, sex, hormone replacement therapy, region, education, height, cancer prior to baseline, water and other healthy beverage intakes, total dietary intakes, physical activity (for diet-only interventions), living alone, measured body weight, number of medications, vitamin, mineral or natural health product consumption, alcohol consumption in the last month, current or former smoker, self-reported diabetes, self-reported high blood pressure, self-reported heart problems, as well as incident hospitalization, bed rest or cancer.

**Figure 3:**
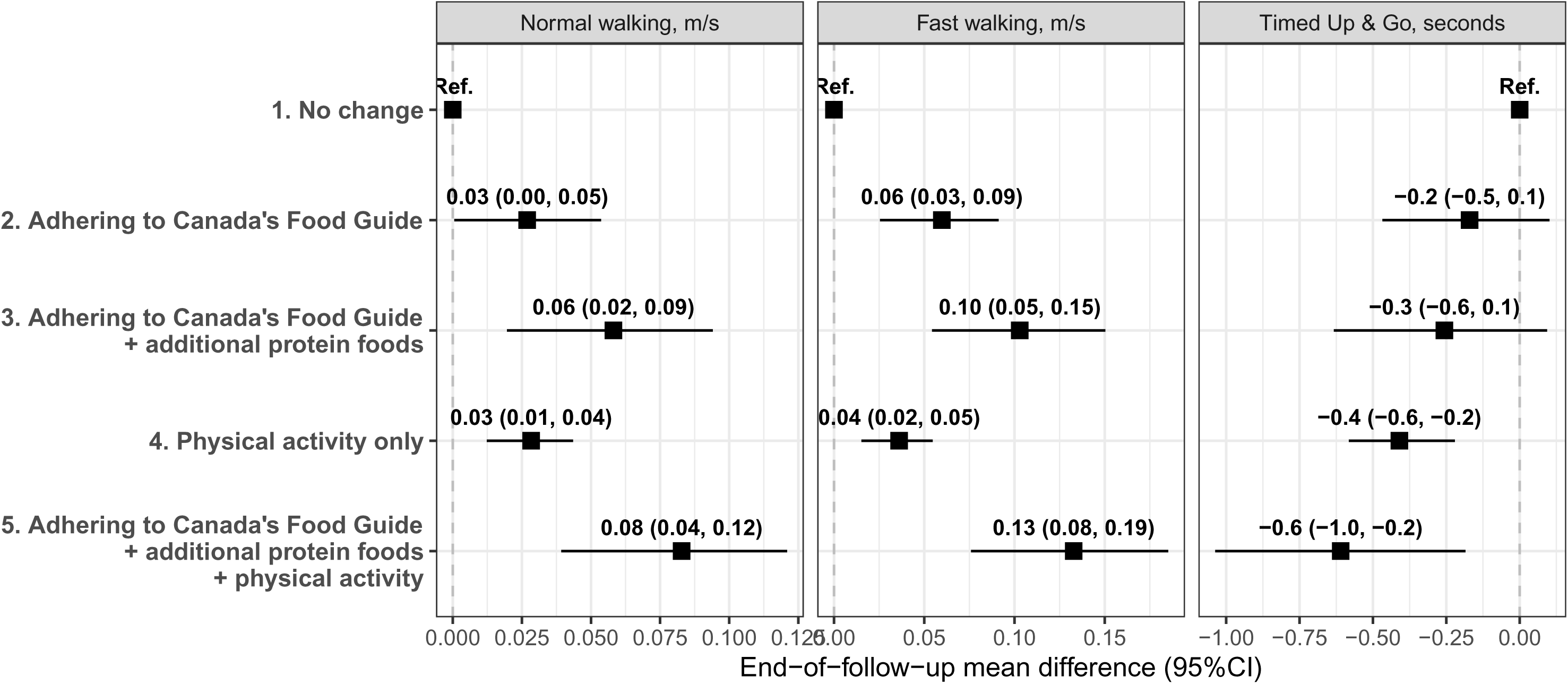
Mean difference in muscle function outcomes after the 3-year hypothetical interventions, compared with no hypothetical intervention, in eligible NuAge participants. Effect estimates were obtained using the g-formula algorithm (see Methods). Physical activity corresponds to that of light intensity or higher. Models were adjusted for the covariates age, sex, hormone replacement therapy, region, education, height, cancer prior to baseline, water and other healthy beverage intakes, total dietary intakes, physical activity (for diet-only interventions), living alone, measured body weight, number of medications, vitamin, mineral or natural health product consumption, alcohol consumption in the last month, current or former smoker, self-reported diabetes, self-reported high blood pressure, self-reported heart problems, as well as incident hospitalization, bed rest or cancer.

**Figure 4:**
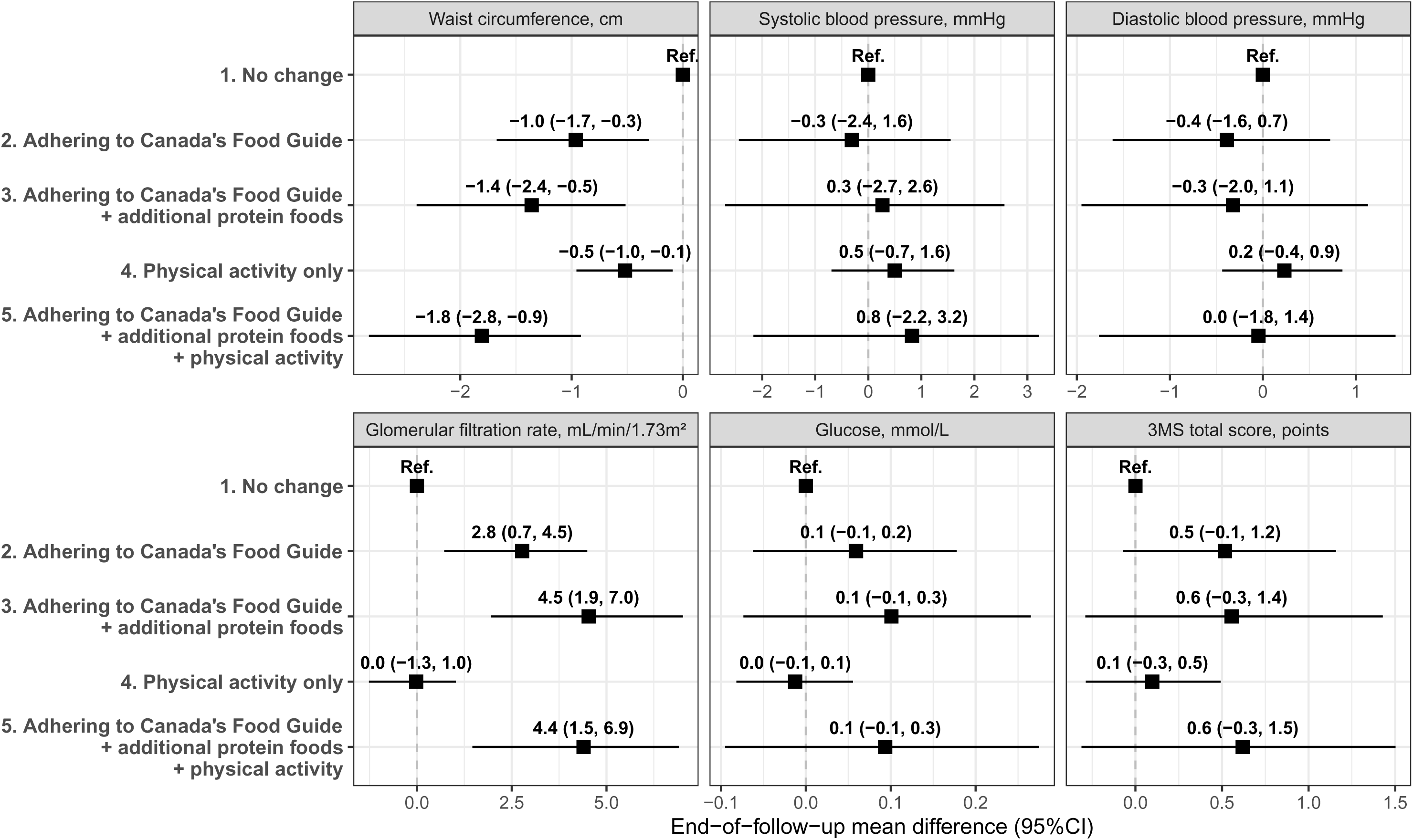
Mean difference in cardiometabolic health and cognition outcomes after the 3-year hypothetical interventions, compared with no hypothetical intervention, in eligible NuAge participants. Effect estimates were obtained using the g-formula algorithm (see Methods). Physical activity corresponds to that of light intensity or higher. Models were adjusted for the covariates age, sex, hormone replacement therapy, region, education, height, cancer prior to baseline, water and other healthy beverage intakes, total dietary intakes, physical activity (for diet-only interventions), living alone, measured body weight, number of medications, vitamin, mineral or natural health product consumption, alcohol consumption in the last month, current or former smoker, self-reported diabetes, self-reported high blood pressure, self-reported heart problems, as well as incident hospitalization, bed rest or cancer.

#### Muscle strength

Compared with no intervention, CFG would have increased elbow flexor strength by 0.8 kg (95%CI: 0.2, 1.3) and quadriceps strength by 0.8 kg (95%CI: 0.0, 1.7). The addition of protein foods did not change these differences. Independently of diet, the PA intervention would have improved elbow flexor by +0.2 kg (95%CI: -0.1, 0.5) and quadriceps strength by +0.9 kg (95%CI: 0.4, 1.4). Thus, adherence to CFG-PLUS resulted in the largest elbow flexor and quadriceps strength increase compared with no intervention (+1.1 kg [95%CI: 0.4, 1.9] and +1.7 kg [95%CI: 0.4, 3.0], respectively). No differences were observed for handgrip strength.

#### Muscle function

All hypothetical interventions would have increased normal and fast walking speed compared with no intervention, but only the inclusion of physical activity improved the Timed Up & Go performance. The Timed Up & Go performance was 0.4 seconds faster (95%CI: -0.6, -0.2) in the PA intervention compared with no intervention. This effect was slightly greater in CFG-PLUS (-0.6 m/s; 95%CI: -1.0, -0.2) compared with no intervention. Similarly, the adherence to the CFG-PLUS intervention would have produced the highest increase in normal walking speed (+0.08 m/s; 95%CI: 0.04, 0.12) and fast walking speed (+0.13 m/s; 95%CI: 0.08, 0.19). More precisely, the increase in normal and fast walking speed were respectively 0.06 m/s (95%CI:0.03, 0.08) and 0.07 m/s (95%CI: 0.04, 0.11) greater in the CFG-PLUS intervention with additional protein foods and physical activity compared with the CFG intervention only.

#### Cardiometabolic health and cognition

Compared with no intervention, all hypothetical interventions reduced waist circumference. The largest reduction was observed for CFG-PLUS (-1.8 cm; 95%CI: - 2.8, -0.9). The estimated glomerular filtration rate was increased after the hypothetical interventions, except for the PA intervention. The largest increase was observed for CFG-PRO (4.5 mL/min/1.73m²; 95%CI: 1.9, 7.0). The CFG intervention was compatible with a small increase for the 3MS total score compared with no intervention (+0.5 points; 95%CI: -0.1, 1.2).

### Sex-stratified analyses

Effect estimates were mostly consistent in their direction but slightly varied in magnitude and precision between males and females (**Figures S2-4)**. For example, compared with no intervention, CFG-PLUS would have increased leg strength by 1.9 kg (95%CI: -0.3, 3.8) in males and 2.2 kg (95%CI: 0.5, 3.8) in females. The estimated effects on eGFR across hypothetical diet interventions was only observed in males (Figure S4).

### Sensitivity analyses

#### Assessment of unmeasured confounding

The 95%CI from the negative control analysis were compatible with both very small decrease and very small increase in albumin concentration, including the null value (Table S10). For example, the adherence to CFG-PLUS was estimated to increase albumin concentration by 0.1 g/L (95%CI -0.4, 0.6) compared with no intervention (Table S10).

#### Assessment of the impact of measurement error

For most outcomes, the biases due to random errors were mostly small on the absolute scale and showed an underestimation of effects (**Table S11**). The biases did not modify the direction of effect estimates, except for elbow flexor strength which showed an effect estimate close to null for the measurement error corrected analysis. The comparison between the error-prone and corrected estimates showed that effects were underestimated by 0.5-fold for quadriceps strength (strength outcomes); underestimated by 0.4-fold for Timed Up & Go and underestimated by 0.7-fold for normal walking (function outcomes); underestimated by 0.3-fold for 3MS total score and underestimated by 0.7-fold for waist circumference (cognition and cardiometabolic health outcomes).

#### Assessment of model misspecification

The g-formula predicted values for time-varying diet, physical activity and covariates were generally consistent with observed values (**Figure S5**).

## Discussion

In this study, we used a target trial emulation to estimate the effects of adhering to CFG recommendations on muscle, cardiometabolic health and cognition in adults aged 67 years and older. We also investigated the value of adding protein foods in combination with physical activity of light intensity or higher in addition to adhering to CFG recommendations. Compared with no hypothetical intervention, we found that adhering to CFG recommendations would have slightly improved arm and leg strength, walking speed, cognitive health score, reduced waist circumference and increased eGFR. The addition of more protein foods and physical activity would have further improved walking speed compared with adherence to CFG recommendations only.

To our knowledge, this study is the first to estimate the impact of adhering to CFG recommendations on muscle function, strength, cardiometabolic health and cognition of older adults. Of note, the hypothetical intervention of adhering to CFG increased protein food intake and overall diet quality (e.g., vegetables and fruits, whole-grain foods) compared with no intervention in this sample. We emphasize that results reflect differences compared with the absence of change in dietary intakes or the amount of physical activity. In other words, effect estimates are not absolute improvements over time but reflect 3-year decline attenuations. For example, we found an end of follow-up increase of 1.7 kg in quadriceps strength in CFG-PLUS compared with no intervention. Accordingly, the observed absolute 3-year change of -2.4 kg under no intervention, corresponding to an annual -3.0% loss, as reported by others (38), would be approximately -0.7 kg under CFG-PLUS (0.9% loss per year) (Table S9). The 1.7 kg-improvement would thus attenuate the observed 3-year decline in leg strength which is a favorable outcome in older adults (38, 39).

To date, no previous RCT have examined the effect of adhering to CFG recommendations per se, alone or in combination with physical activity in older adults. Previous RCT have assessed the effect of various diet intervention, exercise or both on physical frailty, muscle strength or function (40–42). Among nutritional interventions, the effect of protein supplementation, with or without resistance exercise, on muscle strength has been investigated in numerous RCT (43–45).

Over a 2-year follow-up, Teh et al. assessed the effectiveness of a group-based cooking skills and motivation intervention coupled with a strength and balance exercise program on frailty among pre-frail older adults (40). Short-term improvements were found in the combined intervention group, but no meaningful differences in frailty scores persisted at the end of follow-up. Hsieh et al. reported improvements in frailty score, walking speed and leg strength after a 6-month home-based nutrition and exercise interventions, alone or in combination, in older adults (41). Of relevance, the combination of nutrition and exercise intervention yielded the largest difference for walking speed and leg strength compared with both interventions separately (41). In a 4-month RCT, older women experienced 18% greater leg strength gains after resistance exercise when they were provided with 160 g of cooked lean red meat compared with pasta or rice only (42).

Overall, the observation that the inclusion of an exercise or physical activity intervention on top of dietary changes yields the largest gain for walking speed (41) or leg strength (41, 42) is consistent with our findings. The finding of improved strength after increasing the intake of protein is not unanimous. For example, in a meta-analysis of 8 interventions (43), the addition of protein supplements in combination with resistance exercise did not clearly improve leg strength of older adults compared with resistance exercise only (mean difference: +1.4 kg; 95%CI, -1.0 to 3.8). These mixed findings are possibly related to the intervention format (e.g., group vs. individual counselling), intensity, adherence, follow-up duration, and population characteristics. Similarly, considering the quality and bioavailability of protein among different protein foods may also be relevant to optimize muscle strength and function in older adults.

The increase in walking speed compared with no intervention is a noteworthy finding with regard to overall health. Previous literature showed that small meaningful changes in gait speed correspond to a difference of at least 0.03 to 0.05 m/s (46), while a difference up to 0.1 m/s corresponds to substantial improvements (47). A difference of 0.1 m/s in gait speed was also associated with a 12% lower risk of death in a pooled analysis of more than 34,000 community-dwelling older adults aged 65 years or older (48). In the present study, the combination of CFG adherence, additional protein and physical activity yielded gait speed effect estimates (+0.08 m/s; 95%CI: 0.04, 0.12) compatible with substantial improvements and reduced mortality risk. These estimated differences should be interpreted while acknowledging that older adults with walking speed above 1.0 m/s have a low probability of short-term self-reported mobility disability (49), as in the present study (Table S9). Finally, the finding of reduced waist circumference in all hypothetical interventions is also consistent with the reduced 10-y cardiovascular disease risk associated with adherence to CFG recommendations (10).

Strengths of this study include the emulation of a target trial to avoid many fundamental biases of causal effect estimation using non-experimental data, the comprehensive diet and covariate data collection, the ability to examine multiple hypothetical interventions and the sensitivity analyses to assess robustness of findings. Emulating a target trial enabled the analysis of incident rather than prevalent dietary changes, explicit modeling of time-varying intakes (per-protocol contrast), and estimation of sustained intervention effects in all participants rather than extreme group comparisons. Limitations must be highlighted. First, while the 24-h dietary recall is one of the most accurate self-report dietary assessment method, dietary intakes are known to be affected by systematic errors or bias (50, 51) which, in turn, could bias effect estimates unpredictably. Of note, systematic errors contribute less to total error on 24-h dietary recall contrary to other common instruments such as the food frequency questionnaire. Second, most participants had reported dietary intakes and amount of physical activity that did not meet the *a priori* intervention targets (Table S7). Consequently, the effect estimates may be based on a small subset of participants with distinct characteristics, challenging the positivity assumption of causal effect estimation. Third, the NuAge sample size is relatively small and homogenous. Participants are predominantly Caucasians, and the estimated effects may not be generalizable to more diverse populations. Lastly, though sensitivity analyses did not reveal major issues, it does not imply that all assumptions are correct. Our findings hinge on strong assumptions including the lack of unmeasured and residual confounding, which is never guaranteed with observational data.

In conclusion, adhering to CFG 2019 recommendations, compared with no hypothetical intervention, would have improved arm and leg strength as well as walking speed of adults aged 67 years or older over a 3-year follow-up. Increasing the amount of physical activity and the intake protein foods would have yielded clinically-relevant improvements in walking speed compared with CFG alone and no intervention. Our findings support the paradigm of increasing the intake of protein foods and the amount of physical activity in older adults, which should be considered in future studies and dietary guidelines. We stress that these results depend on the assumptions of no unmeasured confounding, no measurement error and correct statistical models. Accordingly, randomized trials are required to minimize these limitations and confirm our results. The target trial described in this study may serve as a starting point for the design of future studies.

## Supporting information

Code snippet

## Posthumous Authorship

Sadly, Nancy Presse passed away in February 2025 during the finalization of this manuscript. We wish to acknowledge her contribution to the present work and to the nutrition and aging research field, in a dedicated career that ended too soon.

## Ethic approval

The NuAge study protocol was approved by the Research Ethics Boards of the Institut universitaire de gériatrie de Montréal and the Institut universitaire de gériatrie de Sherbrooke (Quebec, Canada). The NuAge Database and Biobank have been approved by the Research Ethic Board of the CIUSSS de l’Estrie-CHUS (Quebec, Canada). Ethic approval for secondary analyses of the NuAge Database and Biobank was also obtained from the McGill University Research Ethics Board Office (#22-11-041).

## Data and code availability

Eligible researchers can request access to data from the NuAge Database and Biobank at https://nuage.recherche.usherbrooke.ca/en/demande-dacces/. Final data for the present study were obtained on November 16, 2023. Analysis code will be made available at https://github.com/didierbrassard/NuAge_TargetTrial_CFG

## References

1. Health Canada. Canada’s food guide. Ottawa: Health Canada, 2019,

2. Health Canada. Food, Nutrients and Health: Interim Evidence Update 2018 for Health Professionals and Policy Makers. Ottawa: Health Canada, 2019,

3. Statistics Canada. Table 17-10-0005-01. Population estimates on July 1, by age and gender. 2025. (10.25318/1710000501-eng). (Accessed 2025-12-22 2025).

4. Remond D, Shahar DR, Gille D, et al. Understanding the gastrointestinal tract of the elderly to develop dietary solutions that prevent malnutrition. Oncotarget 2015;6(16):13858–98.

5. Mills CM, Keller HH, DePaul VG, et al. Factors Associated with the Development of High Nutrition Risk: Data from the Canadian Longitudinal Study on Aging. Can J Aging 2023:1–14.

6. Ramage-Morin PL, Garriguet D. Nutritional risk among older Canadians. Health Rep 2013;24(3):3–13.

7. Verlaan S, Ligthart-Melis GC, Wijers SLJ, et al. High Prevalence of Physical Frailty Among Community-Dwelling Malnourished Older Adults-A Systematic Review and Meta-Analysis. J Am Med Dir Assoc 2017;18(5):374–82.

8. Cruz-Jentoft AJ, Sayer AA. Sarcopenia. Lancet 2019;393(10191):2636–46.

9. Wolfe RR. The underappreciated role of muscle in health and disease. Am J Clin Nutr 2006;84(3):475–82.

10. Brassard D, Manikpurage HD, Thériault S, et al. Greater adherence to the 2019 Canada’s Food Guide recommendations on healthy food choices reduces the risk of cardiovascular disease in adults: a prospective analysis of UK Biobank data. Am J Clin Nutr 2022;116(6):1748–58.

11. Cruz-Jentoft AJ, Baeyens JP, Bauer JM, et al. Sarcopenia: European consensus on definition and diagnosis: Report of the European Working Group on Sarcopenia in Older People. Age Ageing 2010;39(4):412–23.

12. Brassard D, Chevalier S. Relationship between adherence to the 2019 Canada’s Food Guide recommendations on healthy food choices and nutrient intakes in older adults. J Nutr 2023;153(9):2699–708.

13. Bauer J, Biolo G, Cederholm T, et al. Evidence-based recommendations for optimal dietary protein intake in older people: a position paper from the PROT-AGE Study Group. J Am Med Dir Assoc 2013;14(8):542–59.

14. Statistics Canada. The Daily — Canadian Health Measures Survey: Activity monitor data, 2018–2019. (https://www150.statcan.gc.ca/n1/daily-quotidien/210901/dq210901c-eng.htm). (Accessed March 2026).

15. Deutz NE, Bauer JM, Barazzoni R, et al. Protein intake and exercise for optimal muscle function with aging: recommendations from the ESPEN Expert Group. Clin Nutr 2014;33(6):929–36.

16. Zeilstra D, Younes JA, Brummer RJ, et al. Perspective: Fundamental Limitations of the Randomized Controlled Trial Method in Nutritional Research: The Example of Probiotics. Adv Nutr 2018;9(5):561–71.

17. Musa-Veloso K, Racey M, MacFarlane A, et al. Challenges in the design, interpretation, and reporting of randomized controlled clinical studies on the health effects of whole foods. Appl Physiol Nutr Metab 2021;46(9):1152–8.

18. Hernan MA, Robins JM. Using Big Data to Emulate a Target Trial When a Randomized Trial Is Not Available. Am J Epidemiol 2016;183(8):758–64.

19. Brassard D, Presse N, Chevalier S. Estimating the Effect of Adhering to the Recommendations of the 2019 Canada’s Food Guide on Health Outcomes in Older Adults: Protocol for a Target Trial Emulation. JMIR Res Protoc 2025:14:e65182.

20. NuAge Database and Biobank. Database and Biobank of the Quebec Longitudinal Study on Nutrition and Successful Aging. 2024. (https://nuage.recherche.usherbrooke.ca). (Accessed Februrary 2024).

21. Gaudreau P, Morais JA, Shatenstein B, et al. Nutrition as a determinant of successful aging: description of the Quebec longitudinal study Nuage and results from cross-sectional pilot studies. Rejuvenation Res 2007;10(3):377–86.

22. Tobias DK, Lajous M. What would the trial be? Emulating randomized dietary intervention trials to estimate causal effects with observational data. Am J Clin Nutr 2021;114(2):416–7.

23. Conway JM, Ingwersen LA, Moshfegh AJ. Accuracy of dietary recall using the USDA five-step multiple-pass method in men: an observational validation study. J Am Diet Assoc 2004;104(4):595–603.

24. Health Canada. Table of Reference Amounts for Food. https://www.canada.ca/en/health-canada/services/technical-documents-labelling-requirements/table-reference-amounts-food.html: Ottawa: Health Canada; 2022. (https://www.canada.ca/en/health-canada/services/technical-documents-labelling-requirements/table-reference-amounts-food.html). (Accessed August 2023).

25. Shatenstein B, Nadon S, Godin C, et al. Development and validation of a food frequency questionnaire. Can J Diet Pract Res 2005;66(2):67–75.

26. Washburn RA, McAuley E, Katula J, et al. The physical activity scale for the elderly (PASE): evidence for validity. J Clin Epidemiol 1999;52(7):643–51.

27. Washburn RA, Smith KW, Jette AM, et al. The Physical Activity Scale for the Elderly (PASE): development and evaluation. J Clin Epidemiol 1993;46(2):153–62.

28. Kipnis V, Freedman LS, Brown CC, et al. Interpretation of energy adjustment models for nutritional epidemiology. Am J Epidemiol 1993;137(12):1376–80.

29. Tomova GD, Arnold KF, Gilthorpe MS, et al. Adjustment for energy intake in nutritional research: a causal inference perspective. Am J Clin Nutr 2022;115(1):189–98.

30. Chiu YH. Well-defined interventions for nutritional studies: from target trials to nutritional modeling. Am J Clin Nutr 2022;115(1):3–5.

31. McGrath S, Lin V, Zhang Z, et al. gfoRmula: An R Package for Estimating the Effects of Sustained Treatment Strategies via the Parametric g-formula. Patterns 2020;1(3).

32. Hernán MA, Robins JM. Causal Inference: What If. CRC Boca Raton: Chapman & Hall/CRC.; 2020.

33. Dickerman BA, Giovannucci E, Pernar CH, et al. Guideline-Based Physical Activity and Survival Among US Men With Nonmetastatic Prostate Cancer. Am J Epidemiol 2019;188(3):579–86.

34. Lipsitch M, Tchetgen Tchetgen E, Cohen T. Negative controls: a tool for detecting confounding and bias in observational studies. Epidemiology 2010;21(3):383–8.

35. Fuhrman MP, Charney P, Mueller CM. Hepatic proteins and nutrition assessment. J Am Diet Assoc 2004;104(8):1258–64.

36. Zhang S, Midthune D, Guenther PM, et al. A New Multivariate Measurement Error Model with Zero-Inflated Dietary Data, and Its Application to Dietary Assessment. Ann Appl Stat 2011;5(2B):1456–87.

37. Chiu YH, Wen L, McGrath S, et al. Evaluating Model Specification When Using the Parametric G-Formula in the Presence of Censoring. Am J Epidemiol 2023;192(11):1887–95.

38. Goodpaster BH, Park SW, Harris TB, et al. The loss of skeletal muscle strength, mass, and quality in older adults: the health, aging and body composition study. J Gerontol A Biol Sci Med Sci 2006;61(10):1059–64.

39. Ferrucci L, de Cabo R, Knuth ND, et al. Of Greek heroes, wiggling worms, mighty mice, and old body builders. J Gerontol A Biol Sci Med Sci 2012;67(1):13–6.

40. Teh R, Barnett D, Edlin R, et al. Effectiveness of a complex intervention of group-based nutrition and physical activity to prevent frailty in pre-frail older adults (SUPER): a randomised controlled trial. Lancet Healthy Longev 2022;3(8):e519–e30.

41. Hsieh TJ, Su SC, Chen CW, et al. Individualized home-based exercise and nutrition interventions improve frailty in older adults: a randomized controlled trial. Int J Behav Nutr Phys Act 2019;16(1):119.

42. Daly RM, O’Connell SL, Mundell NL, et al. Protein-enriched diet, with the use of lean red meat, combined with progressive resistance training enhances lean tissue mass and muscle strength and reduces circulating IL-6 concentrations in elderly women: a cluster randomized controlled trial. Am J Clin Nutr 2014;99(4):899–910.

43. Kirwan RP, Mazidi M, Rodriguez Garcia C, et al. Protein interventions augment the effect of resistance exercise on appendicular lean mass and handgrip strength in older adults: a systematic review and meta-analysis of randomized controlled trials. Am J Clin Nutr 2022;115(3):897–913.

44. Mertz KH, Reitelseder S, Bechshoeft R, et al. The effect of daily protein supplementation, with or without resistance training for 1 year, on muscle size, strength, and function in healthy older adults: A randomized controlled trial. Am J Clin Nutr 2021;113(4):790–800.

45. Whaikid P, Piaseu N. The effectiveness of protein supplementation combined with resistance exercise programs among community-dwelling older adults with sarcopenia: a systematic review and meta-analysis. Epidemiol Health 2024;46:e2024030.

46. Kwon S, Perera S, Pahor M, et al. What is a meaningful change in physical performance? Findings from a clinical trial in older adults (the LIFE-P study). J Nutr Health Aging 2009;13(6):538–44.

47. Perera S, Mody SH, Woodman RC, et al. Meaningful change and responsiveness in common physical performance measures in older adults. J Am Geriatr Soc 2006;54(5):743–9.

48. Studenski S, Perera S, Patel K, et al. Gait speed and survival in older adults. JAMA 2011;305(1):50–8.

49. Miller ME, Magaziner J, Marsh AP, et al. Gait Speed and Mobility Disability: Revisiting Meaningful Levels in Diverse Clinical Populations. J Am Geriatr Soc 2018;66(5):954–61.

50. Freedman LS, Commins JM, Moler JE, et al. Pooled results from 5 validation studies of dietary self-report instruments using recovery biomarkers for potassium and sodium intake. Am J Epidemiol 2015;181(7):473–87.

51. Freedman LS, Commins JM, Moler JE, et al. Pooled results from 5 validation studies of dietary self-report instruments using recovery biomarkers for energy and protein intake. Am J Epidemiol 2014;180(2):172–88.

