## Supplemental Material for "A target trial emulation of adherence to Canada’s Food Guide 2019 recommendations in older adults"

### Supplemental Methods

#### Study Design

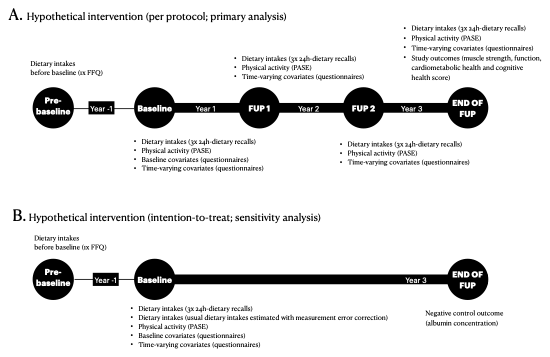

**Figure S1**. Schematic overview of the study design using the NuAge Biobank and Database. FFQ, food-frequency questionnaire; FUP, follow-up; NuAge, Quebec Longitudinal Study on Nutrition and Successful Aging. PASE, Physical Activity Scale for the Elderly.

#### Hypothetical intervention

Table S1 presents the detailed hypothetical diet and physical activity intervention. As a threshold intervention, participants would be expected to meet or exceed the target dietary intakes or amount of physical activity per day at each follow-up. Complete details about the hypothetical intervention are available elsewhere (Brassard, Presse, and Chevalier 2025).

**Table S1.** Hypothetical diet and physical activity interventions emulated in the NuAge study, by sex^1^

|  | **Recommended foods, RA/day** | | | | | | **Foods and beverages not recommended, RA/day*^2^*** | | |  |
| --- | --- | --- | --- | --- | --- | --- | --- | --- | --- | --- |
| **Sex** | Vegetables & fruits | Whole grains | Protein foods, plant-based | Protein foods, animal-based | Milk & Plant-based bev. with protein | Unsaturated oils & fats | Other foods | Sugary drinks, alcohol | Non-whole grains | Physical activity, minutes/day*^3^* |
| **1. Natural course (no intervention)*^4^*** | | | | | | | | | | |
| Males | 4.4 (2.1) | 1.0 (0.9) | 0.6 (0.7) | 2.5 (1.3) | 0.7 (0.8) | 1.1 (1.2) | 4.9 (3.6) | 1.3 (1.2) | 1.8 (1.1) | 15.0 (0.0, 45.0) |
| Females | 4.5 (1.9) | 0.8 (0.6) | 0.5 (0.6) | 2.2 (1.1) | 0.7 (0.7) | 1.0 (0.9) | 3.5 (2.4) | 0.8 (0.7) | 1.5 (0.9) | 6.4 (0.0, 30.0) |
| **2. Adhering to Canada's Food Guide 2019 recommendations on healthy food choices*^5^*** | | | | | | | | | | |
| Males | 6 | 1.5 | 1 | 2 | 1 | 1 | Proportionately lower than what is required to meet the intervention threshold of foods recommended | | | No intervention |
| Females | 5 | 1.5 | 0.8 | 1.5 | 1 | 1 | Proportionately lower than what is required to meet the intervention threshold of foods recommended | | | No intervention |
| **3. Same as 2 + additional protein foods*^6^*** | | | | | | | | | | |
| Males | 6 | 1.5 | 1.5 | 3.5 | 1.5 | 1 | Proportionately lower than what is required to meet the intervention threshold of foods recommended | | | No intervention |
| Females | 5 | 1.5 | 1.3 | 3 | 1.5 | 1 | Proportionately lower than what is required to meet the intervention threshold of foods recommended | | | No intervention |
| **4. Physical activity only** | | | | | | | | | | |
| Males | No intervention | | | | | | | | | 30 or more |
| Females | No intervention | | | | | | | | | 30 or more |
| **5. Same as 2 + additional protein foods + physical activity** | | | | | | | | | | |
| Males | 6 | 1.5 | 1.5 | 3.5 | 1.5 | 1 | Proportionately lower than what is required to meet the intervention threshold of foods recommended | | | 30 or more |
| Females | 5 | 1.5 | 1.3 | 3 | 1.5 | 1 | Proportionately lower than what is required to meet the intervention threshold of foods recommended | | | 30 or more |
| *^1^*As a threshold intervention, participants would be expected to meet or exceed the target dietary intakes of recommend foods or amount of physical activity per day at each follow-up. CFG, Canada's Food Guide; NA, not available; NuAge, Quebec Longitudinal Study on Nutrition and Successful Aging; RA, reference amount; y, year | | | | | | | | | | |
| *^2^* The target trial would be an isocaloric intervention. The target trial emulation analysis adjusts for total food and beverage intakes. Consumption of **foods not recommended** is thus set at the minimal level that permits a concomitant increase in recommended foods to meet Canada's Food Guide targets. In other words, the number of reference amounts for **foods not recommended** varies across individuals according to the extent to which their reported intakes fall below the targets for recommended foods. | | | | | | | | | | |
| *^3^*For reference purpose, values are median (interquartile range) amount of physical activity observed at baseline among eligible NuAge participants. Physical activity corresponds to that of light intensity or higher. Participants would be expected to meet or exceed the target amount of physical activity per day. | | | | | | | | | | |
| *^4^*For reference purpose, values are mean (SD) dietary intakes at baseline among eligible NuAge participants. In other words, values are the observed intakes for the food categories or amount of physical activity when no intervention is applied. Dietary intakes were calculated using the mean of data from all 24-h dietary recalls completed at baseline. | | | | | | | | | | |
| *^5^*Values are derived from Health Canada's simulated composite diets of adults 71 years or older. Participants would be expected to meet or exceed these targets for each food categories. The specific food choices within these categories would be at the participants' discretion. Values for vegetables and fruits, whole-grain foods and plant-based protein foods were truncated to correspond, at most, to the 90th percentile of the distribution of usual intakes among Canadians aged 65 years in 2015. | | | | | | | | | | |
| *^6^*Extra protein foods were added as follows: +0.5 RA of plant-based protein foods (e.g., 25 grams of nuts), +1.5 RA of animal-based protein foods (e.g., 150 grams of cooked unprocessed red meat, fish or poultry or 3 small eggs), +0.5 RA of milk or plant-based beverage with protein (e.g., 125 ml of milk or plant-based beverages with sufficient protein). | | | | | | | | | | |

#### Exposure: diet via 24-h dietary recalls

Reported foods and beverages were mapped to the Canadian Nutrient File 2007b to derive nutrient intakes, which corresponds to the nutritional value of foods and beverages available at the time of data collection. Nutritional supplements consumption was assessed using the 24-h dietary recalls, while vitamins, minerals or natural health product consumption was assessed during interviews (see below for covariates).

To reduce the influence of extreme values in the hypothetical intervention modelling, the upper values of average dietary intakes reported using 24-h dietary recall were winsorized at the value corresponding to the upper quartile plus twice the interquartile range. For descriptive purposes, the overall adherence to CFG’s recommendations at baseline was estimated using the HEFI-2019 score (Brassard, Elvidge Munene, St-Pierre, Guenther, et al. 2022; Brassard, Elvidge Munene, St-Pierre, Gonzalez, et al. 2022). HEFI-2019 scores were based on the average intake data from all 24-h dietary recalls completed at baseline. The HEFI-2019 score ranges from 0 to 80 points. Higher scores indicate greater adherence to CFG recommendations on healthy food choices (Brassard, Elvidge Munene, St-Pierre, Guenther, et al. 2022).

#### Outcome measurements

**Table S2**. Outcome and measurement procedures in the NuAge study

| **Outcomes** | **NuAge variable name** | **Description and measurement** | **Reference** |
| --- | --- | --- | --- |
| **Muscle strength** | | | |
| Handgrip strength, kPa | MAXVD0, MAXVG0 | Handgrip strength was measured using a pneumatic dynamometer (Martin vigorimeter). Three maximum contractions were recorded on each hand. The highest value in all attempts was used. | Payette et al. (1998) |
| Elbow flexor strength, kg | MAXBD0, MAXBG0 | The maximum voluntary isometric strength of the elbow flexors was measured using a dynamometer Microfet2 (Hoggan Industries, Inc.) and the belt-resisted method. | Payette et al. (1998) |
| Knee extensors (quadriceps) strength, kg | MAXQD0, MAXQG0 | The maximum voluntary isometric strength of knee extensors was measured using a dynamometer Microfet2 (Hoggan Industries, Inc.) and the belt-resisted method. | Desrosiers et al. (1998) |
| **Muscle function** | | | |
| Normal walking, m/s | MARCH1, MARCH2 | Participants were asked to walk at their usual pace over a 4-meters course twice. The best time was recorded. | — |
| Fast walking, m/s | MARRAP | Same as normal walking, but participants were asked to walk as fast as they could. | — |
| Timed Up & Go, seconds | TUG | Time required to rise from a chair, walk 3 meters, turn around, walk back, and sit down. | Podsiadlo and Richardson (1991) |
| **Cardiometabolic health** | | | |
| Waist circumference, cm | CIRCTA | Measured using a standard tape (fiberglass tape) | — |
| Systolic blood pressure, mmHg | TAMAX | Sitting blood pressure | — |
| Diastolic blood pressure, mmHg | TAMIN | Sitting blood pressure | — |
| Glucose, mmol/L | GLUCOS | Blood samples were collected after an overnight fast. Glucose concentration assessed with fresh blood samples [clinical biochemistry laboratories of St. Mary's Hospital (Montreal site) and Sherbrooke University Health Centre (Sherbrooke site)] | — |
| Glomerular filtration rate, mL/min/1.73m^2 | GFR | Calculated using serum creatinine concentration | — |
| **Cognitive health** | | | |
| Modified Mini-Mental State Examination (3MS) score | SCOTMS, VALTMS | Total 3MS score | Teng and Chui (1987) |
| **Negative control** | | | |
| Albumin, g/L | ALBUMI | Blood samples were collected after an overnight fast. Albumin concentration assessed with fresh blood samples [clinical biochemistry laboratories of St. Mary's Hospital (Montreal site) and Sherbrooke University Health Centre (Sherbrooke site)] | — |

#### Statistical analysis

##### Sample Size

For a target trial emulation study, *a priori* power analyses are unnecessary (Hernán 2021). Thus, the final sample size was determined by including data from all eligible participants in the NuAge Database and Biobank.

##### G-formula algorithm

The g-formula algorithm involves multiple steps (McGrath et al. 2020; Hernán and Robins 2020). First, the predicted values of each time-varying variables (including both covariates and intervention variables) were estimated using separate regression models. Each model predicted the values throughout follow-up of a given time-varying variable while adjusting for the history of intervention (diet and physical activity) and covariates, as well as adjusting for baseline covariates and prior dietary intakes (where applicable). Second, copies of the data were generated using Monte Carlo simulations. In these copies, the value of each intervention variables is modified according to the pre-specified hypothetical interventions. For example, to simulate the intervention “adhering to CFG” in one copy of the data, the values of the dietary components are increased to ensure that all individuals meet the pre-specified thresholds as detailed in Table S1. Third, the regression coefficients estimated in the first step were then applied to the modified Monte Carlo simulations data to estimate the counterfactual, unobserved, value of the outcome at the end of follow-up. This value corresponds to the outcome, had everyone adhered to the intervention at each of the four follow-ups (denoted $Y^{X=1,1,1,1}$). Fourth, the counterfactual outcome was compared with the observed outcome under the natural course (denoted $Y^{\left( X=0,0,0,0 \right)}$), i.e., when no changes to the intervention variable were made. The difference between the counterfactual and natural course outcome values correspond to the estimated causal effect of the hypothetical interventions, i.e. $E\left( Y^{X=1,1,1,1} \right)-E\left( Y^{X=0,0,0,0} \right)$ (Brassard, Presse, and Chevalier 2025). Fifth, steps 1 to 4 are repeated across 500 bootstrap resamples to estimate variance using the percentile method. Table S3 describes the modelling approach for each variable.

Of note, intervention variables of diet and physical activity were not dichotomized prior to modelling. Rather, the regression coefficient of each variable was estimated in step 1 using flexible modelling as presented in Table S3. In step 2, only the intervention variables in the simulated data generated were modified (after modelling). Finally, in step 3, the outcome model is applied to the simulated data to obtain expected values of outcome under each of the hypothetical intervention.

##### Detailed variable modeling

Table S3 shows how each variable was modeled in the g-formula algorithm.

**Table S3.** Modeling approach for each variable in the g-formula algorithm

| **Variable** | **Modeling as independent variable** | **Time-varying variable***^1^* | **Modeling as dependent variable** |
| --- | --- | --- | --- |
| **Covariates** | | | |
| Age | Continuous: restricted cubic spline with 3 knots | No | Not applicable |
| Sex | Dichotomous | No | Not applicable |
| Hormone replacement therapy | Dichotomous | No | Not applicable |
| Region | Categorical, 3 levels | No | Not applicable |
| Education | Continuous: restricted cubic spline with 3 knots | No | Not applicable |
| Height | Continuous: restricted cubic spline with 5 knots | No | Not applicable |
| Cancer prior to baseline | Dichotomous | No | Not applicable |
| Diet: Water and other healthy beverage intakes | Continuous: linear | Yes | Normal (linear regression) |
| Diet: Total dietary intakes | Continuous: restricted cubic spline with 4 knots | Yes | Normal (linear regression) |
| Living alone | Dichotomous | Yes | Binomial (logistic regression) |
| Measured body weight | Continuous: restricted cubic spline with 5 knots | Yes | Normal (linear regression) |
| Number of medications | Continuous: restricted cubic spline with 3 knots | Yes | Normal (linear regression) |
| Vitamin, mineral or natural health product consumption | Dichotomous | Yes | Binomial (logistic regression) |
| Alcohol consumption in the last month | Dichotomous | Yes | Binomial (logistic regression) |
| Current or former smoker | Dichotomous | Yes | Binomial (logistic regression) |
| Self-reported diabetes | Dichotomous | Yes | Binomial (logistic regression) |
| Self-reported high blood pressure | Dichotomous | Yes | Binomial (logistic regression) |
| Self-reported heart problems | Dichotomous | Yes | Binomial (logistic regression) |
| Incident hospitalization, bed rest or cancer | Dichotomous | Yes | Binomial (logistic regression) |
| **Intervention variables** | | | |
| Vegetables and fruits | Continuous: linear | Yes | Normal (linear regression) |
| Whole-grain foods | Continuous: linear | Yes | Normal (linear regression) |
| Plant-based protein foods | Continuous: restricted cubic spline with 3 knots | Yes | Zero-inflated normal (logistic then log-linear regression) |
| Animal-based protein foods | Continuous: restricted cubic spline with 3 knots | Yes | Normal (linear regression) |
| Unsweetened milk | Continuous: restricted cubic spline with 3 knots | Yes | Normal (linear regression) |
| Unsaturated fats and oils | Continuous: linear | Yes | Zero-inflated normal (logistic then log-linear regression) |
| Light or higher intensity physical activities | Continuous: restricted cubic spline with 4 knots | Yes | Zero-inflated normal (logistic then log-linear regression) |
| *^1^* Data on time-varying variables were collected at each annual follow-up over 3 years (baseline, follow-up 1, follow-2, end of follow-up) | | | |

##### Missing time-varying diet, physical activity and covariate data

**Table S4**. Proportion of missing time-varying diet, physical activity and covariates across follow-ups in eligible NuAge participants^1^

| **Characteristic** | **Baseline** N = 1,561 | **Follow-up 1** N = 1,488 | **Follow-up 2** N = 1,390 | **End of follow-up** N = 1,285 |
| --- | --- | --- | --- | --- |
| Dietary intakes (24-h dietary recall) | 0 (0%) | 68 (4.6%) | 37 (2.7%) | 58 (4.5%) |
| Total light or higher intensity activities | 0 (0%) | 104 (7.0%) | 76 (5.5%) | 148 (12%) |
| Living alone | 0 (0%) | 95 (6.4%) | 52 (3.7%) | 95 (7.4%) |
| Measured body weight | 0 (0%) | 106 (7.1%) | 46 (3.3%) | 59 (4.6%) |
| Number of medications | 0 (0%) | 87 (5.8%) | 31 (2.2%) | 45 (3.5%) |
| Vitamin, mineral or natural health product consumption | 0 (0%) | 87 (5.8%) | 31 (2.2%) | 45 (3.5%) |
| Alcohol consumption in the last month | 0 (0%) | 96 (6.5%) | 52 (3.7%) | 96 (7.5%) |
| Smoking status | 0 (0%) | 95 (6.4%) | 52 (3.7%) | 95 (7.4%) |
| Has diabetes | 0 (0%) | 95 (6.4%) | 52 (3.7%) | 95 (7.4%) |
| Has high blood pressure | 0 (0%) | 95 (6.4%) | 52 (3.7%) | 95 (7.4%) |
| Has heart problems | 0 (0%) | 95 (6.4%) | 52 (3.7%) | 95 (7.4%) |
| *^1^*Values are n (%). When data missing were missing at one follow-up, the value from the previous follow-up was carried forward once. Participants were considered as lost to follow-up if questionnaire non-response occurred at 2 consecutive follow-ups (see Methods). As per eligibility criteria, there were no missing values for time-varying covariate at baseline or time-invariant covariates (not shown: age, sex, hormone replacement therapy, region, education, height, dietary intakes via food-frequency questionnaire). | | | | |

##### Sensitivity analysis to assess the impact of measurement error

Dietary intake data from short-term instruments, including 24-h dietary recalls, are mainly affected by within-individual random measurement error (Thompson et al. 2015). Variables with correlated random errors can bias effect estimates in any direction in a model (Keogh et al. 2020). The impact of random errors associated with 24-h dietary recalls data was assessed by comparing the uncorrected effect estimates with corrected effect estimates (Brassard, Presse, and Chevalier 2025).

Data from all 24-h dietary recalls completed at baseline were used to apply the National Cancer Institute (NCI) Markov Chain Monte Carlo (MCMC) multivariate method (Zhang et al. 2011). The NCI multivariate method allows obtaining measurement error-corrected estimates using regression calibration (Zhang et al. 2011). For the present analysis, key assumptions are that 24-h dietary recalls data are unbiased (i.e., no systematic error) at the participant level and that day-to-day variations are generalizable to the complete sample (Kirkpatrick et al. 2022). Of note, the NCI method has not yet been extent to time-varying dietary intakes as in the NuAge study. Thus, the analysis described below corresponds to an observational analog of the intention-to-treat causal contrast where the hypothetical intervention is assigned at baseline only and non-adherence is not considered (Brassard, Presse, and Chevalier 2025). In other words, only the baseline data was used for the sensitivity analysis.

The NCI multivariate method involves multiple steps. First, the best box-cox transformations to approximate a normal distribution were estimated for all dietary constituents ($X_{j}$). The transformation was conditional on all other model covariates, i.e., pre-baseline dietary intakes and baseline covariates ($L$). Then, dietary constituents were separated according to their pattern of consumption, either daily or episodic. In the present study, dietary constituents were classified as episodic if they were not reported by >10% of participants on the first 24-h dietary recall. The 5 daily foods (proportion of zeros) were non-whole grain foods (9.4%), other low nutritive value foods (4.6%), animal-based protein foods (3.1%), vegetables and fruits (1.5%), and water and other healthy beverages (1.2%), and the 6 episodic foods were saturated fats and oils (58.8%), plant-based protein foods (52.4%), whole-grain foods (29.2%), unsaturated fats and oils (27.5%), sugary drinks and alcohol (26.3%), and unsweetened milk or plant-based beverages (16.9%). Second, the NCI multivariate measurement model estimated and "removed random errors" while accounting for the correlation between all dietary constituents and their errors, the probability and amount of consumption as well as considering the outcome model covariates. Third, 500 conditional draws per participant were generated and used in the Monte Carlo simulation step of the NCI method, thus yielding 500 simulations based on observed data for each participant. Using the complete simulation data, all derived variables were calculated. New variables included total intakes (e.g., total food intakes) and restricted cubic spline transformations, where applicable (i.e., for animal-based protein foods, plant-based protein foods, unsweetened milk and total dietary intakes; see Table S3). Copies of the data were also generated at this step, denoted as $X_{j}=0$ for no intervention (natural course) and $X_{j}=1$ for the hypothetical CFG intervention. The dietary intake data values for the hypothetical intervention components were modified to meet the intended targets (see the intervention *Adhering to Canada’s Food Guide 2019 recommendations on healthy food choices* in Table S1). Fourth, simulated data were averaged at the participant and intervention level to obtain the best conditional expectations of "usual intakes", given on observed data for each participant, to be used in the regression calibration procedure. Fifth, multivariable linear regression models and standardization (Hernán and Robins 2020) were used to estimate end of follow-up outcome values $Y$, given intervention variables $X$ and covariates $L$. That is, $E\left[ Y|X_{j}=0,L=l \right]$ for no intervention (natural course) and $E\left[ Y|X_{j}=1,L=l \right]$ for Canada’s Food Guide intervention. The measurement-error corrected effect of the CFG intervention, $\theta$, was calculated as the difference between the outcome value under the hypothetical intervention and the outcome value under no intervention.

Finally, the same analysis was repeated using unmodeled dietary intake data at baseline to obtain measurement error-uncorrected effect estimates ($\hat{\theta}$). Ratios ($\hat{\theta}/\theta$) and differences ($\hat{\theta}-\theta$) were calculated to estimate the relative and absolute impact of impact of random measurement error, respectively.

### Supplemental Results

#### Characteristics of participants by eligibility

**Table S5**. Characteristics of NuAge participants according to their baseline eligibility for the target trial emulation^1^

| **Characteristic** | **Eligible** N = 1,561*^2^* | **Non-eligible** N = 192*^2^* | **p-value***^3^* |
| --- | --- | --- | --- |
| Age, years | 74 (4) | 75 (4) | <0.001 |
| Females | 809 (52%) | 104 (54%) | 0.5 |
| Region |  |  | <0.001 |
| Sherbrooke | 799 (51%) | 119 (62%) |  |
| Montréal | 448 (29%) | 55 (29%) |  |
| Laval | 314 (20%) | 18 (9.4%) |  |
| Education, years | 12 (4) | 11 (4) | 0.005 |
| Household income, $CAD | 33,100 [26,900] | 30,000 [20,525] | 0.001 |
| (Missing) | 201 | 28 |  |
| Marital status |  |  | 0.003 |
| Single | 167 (11%) | 22 (11%) |  |
| Widowed | 323 (21%) | 60 (31%) |  |
| Married | 944 (60%) | 92 (48%) |  |
| Separated, divorced | 127 (8.1%) | 18 (9.4%) |  |
| Living alone | 494 (32%) | 80 (42%) | 0.005 |
| Ethnicity |  |  | 0.2 |
| Caucasian | 1,540 (99%) | 187 (97%) |  |
| Other | 21 (1.3%) | 5 (2.6%) |  |
| Body mass index, kg/m² | 27.8 (4.4) | 28.1 (4.8) | 0.3 |
| (Missing) | 0 | 6 |  |
| Body mass index category |  |  | 0.2 |
| Healthy | 416 (27%) | 52 (28%) |  |
| Overweight | 743 (48%) | 76 (41%) |  |
| Obesity | 402 (26%) | 58 (31%) |  |
| (Missing) | 0 | 6 |  |
| Total light or higher intensity activities, minutes/day | 13 [39] | 0 [26] | 0.14 |
| (Missing) | 0 | 5 |  |
| Meeting physical activity guidelines (150 minutes/week) | 404 (26%) | 46 (25%) | 0.7 |
| (Missing) | 0 | 5 |  |
| Smoking status |  |  | 0.6 |
| No | 809 (52%) | 107 (56%) |  |
| Formerly | 647 (41%) | 72 (38%) |  |
| Yes | 105 (6.7%) | 13 (6.8%) |  |
| Alcohol consumption in the last month | 1,265 (81%) | 143 (74%) | 0.031 |
| Vitamin, mineral or natural health product consumption | 492 (32%) | 60 (31%) | >0.9 |
| Difficulty chewing or meal interruption due to mouth or teeth problems | 301 (19%) | 38 (20%) | 0.8 |
| (Missing) | 0 | 1 |  |
| Self-reported high blood pressure | 733 (47%) | 97 (51%) | 0.4 |
| Self-reported diabetes | 165 (11%) | 30 (16%) | 0.036 |
| Self-reported heart problems | 342 (22%) | 47 (24%) | 0.4 |
| Number of medications | 4 [5] | 5 [3] | 0.3 |
| Past or current hormone replacement therapy (females only) | 451 (56%) | 51 (49%) | 0.2 |
| (Missing) | 752 | 88 |  |
| Total Healthy Eating Food Index (/80)*^4^* | 47 (9) | 46 (10) | 0.14 |
| (Missing) | 0 | 8 |  |
| Number of 24-h dietary recalls completed at baseline |  |  | <0.001 |
| 0 | 0 (0%) | 8 (4.2%) |  |
| 1 | 4 (0.3%) | 10 (5.2%) |  |
| 2 | 30 (1.9%) | 4 (2.1%) |  |
| 3 | 1,527 (98%) | 170 (89%) |  |
| *^1^*Eligibility criteria at baseline for the target trial emulation were to have no missing data and to have valid dietary intakes data (at least one 24-hour dietary recall with 500 kcal or more). | | | |
| *^2^*Mean (SD); n (%); Median [IQR] | | | |
| *^3^*Wilcoxon rank sum test; Pearson's Chi-squared test; Fisher's exact test | | | |
| *^4^*The total Healthy Eating Food Index (HEFI)-2019 total score reflects overall adherence to Canda's Food Guide recommendations on healthy food choices (range 0-80). Mean scores were calculated using the simple method. Dietary intakes for the scoring algorithm were based on the average intakes among all 24-h dietary recalls completed at baseline by each participant. | | | |

#### Observed dietary intakes

***Table S6***: Observed values of dietary intakes and physical activity per day across follow-ups in eligible NuAge participants (n=1,561)*^1^*

| **Characteristic** | **Baseline** | **Follow-up 1** | **Follow-up 2** | **End of follow-up** |
| --- | --- | --- | --- | --- |
| Vegetables and fruits | 4.4 (2.0) | 4.5 (2.1) | 4.3 (2.0) | 4.3 (2.0) |
| Whole-grain foods | 0.9 (0.8) | 0.9 (0.8) | 0.9 (0.7) | 0.8 (0.7) |
| Protein foods, plant-based | 0.5 (0.7) | 0.5 (0.7) | 0.5 (0.7) | 0.5 (0.7) |
| Protein foods, animal-based | 2.4 (1.2) | 2.3 (1.1) | 2.3 (1.3) | 2.2 (1.2) |
| Milk, unsweetened | 0.7 (0.8) | 0.7 (0.8) | 0.7 (0.7) | 0.7 (0.7) |
| Fats and oils, unsaturated | 1.1 (1.0) | 1.0 (1.1) | 1.0 (1.0) | 0.9 (1.0) |
| Other foods | 4.2 (3.1) | 4.3 (2.9) | 4.3 (3.2) | 4.3 (3.2) |
| Other beverages | 1.1 (1.0) | 1.1 (1.1) | 1.1 (1.0) | 1.0 (1.0) |
| Non-whole grain foods | 1.7 (1.0) | 1.6 (1.1) | 1.7 (1.0) | 1.6 (1.0) |
| Light or higher intensity physical activities, minutes/day | 12.9 (0.0, 38.6) | 0.0 (0.0, 25.7) | 0.0 (0.0, 25.7) | 6.4 (0.0, 25.7) |
| *^1^* Data in this table exclude values carried forward from previous follow-ups. For dietary intakes, values are mean (SD) of data from all 24-h dietary recalls completed at each follow-up. Values are expressed in reference amounts. A total of 0, 139, 188 and 286 participants had missing values at baseline, follow-up 1, follow-up 2 and end of follow-up, respectively. For physical activity, values are median (Q1, Q3). A total of 0, 177, 242 and 408 participants had missing values at baseline, follow-up 1, follow-up 2 and end of follow-up, respectively. | | | | |

#### Proportion of participants meeting the hypothetical intervention thresholds

***Table S7***: Observed proportions of participants meeting intervention thresholds across follow-up in eligible NuAge participants (n=1,561)*^1^*

| **Intervention component label** | **Baseline** | **Follow-up 1** | **Follow-up 2** | **End of follow-up** |
| --- | --- | --- | --- | --- |
| **Adhering to Canada's Food Guide – Males (n=752)** | | | | |
| Vegetables and fruits ≥ 6 RA/day | 148 (19.7%) | 140 (20.4%) | 118 (18.1%) | 117 (19.2%) |
| Whole-grain foods ≥ 1.5 RA/day | 161 (21.4%) | 156 (22.8%) | 133 (20.4%) | 108 (17.8%) |
| Protein foods, plant-based ≥ 1 RA/day | 167 (22.2%) | 146 (21.3%) | 152 (23.3%) | 141 (23.2%) |
| Protein foods, animal-based ≥ 2 RA/day | 447 (59.4%) | 385 (56.2%) | 352 (53.9%) | 332 (54.6%) |
| Milk and plant-based bev. with protein ≥ 1 RA/day | 202 (26.9%) | 183 (26.7%) | 172 (26.3%) | 161 (26.5%) |
| Unsaturated oils and fats ≥ 1 RA/day | 328 (43.6%) | 284 (41.5%) | 265 (40.6%) | 226 (37.2%) |
| **Adhering to Canada's Food Guide + additional protein foods – Males** | | | | |
| Protein foods, plant-based ≥ 1.5 RA/day | 88 (11.7%) | 88 (12.8%) | 79 (12.1%) | 87 (14.3%) |
| Protein foods, animal-based ≥ 3.5 RA/day | 135 (18.0%) | 102 (14.9%) | 105 (16.1%) | 100 (16.4%) |
| Milk and plant-based bev. with protein ≥ 1.5 RA/day | 87 (11.6%) | 83 (12.1%) | 76 (11.6%) | 67 (11.0%) |
| **Physical activity only – Males** | | | | |
| Physical activity ≥ 30 minutes/day | 258 (34.3%) | 192 (28.3%) | 179 (28.1%) | 157 (27.9%) |
| **Adhering to Canada's Food Guide – Females (n=809)** | | | | |
| Vegetables and fruits ≥ 5 RA/day | 280 (34.6%) | 270 (36.6%) | 231 (32.1%) | 215 (32.2%) |
| Whole-grain foods ≥ 1.5 RA/day | 118 (14.6%) | 103 (14.0%) | 90 (12.5%) | 89 (13.3%) |
| Protein foods, plant-based ≥ 0.8 RA/day | 159 (19.7%) | 143 (19.4%) | 146 (20.3%) | 133 (19.9%) |
| Protein foods, animal-based ≥ 1.5 RA/day | 588 (72.7%) | 534 (72.5%) | 500 (69.4%) | 456 (68.4%) |
| Milk and plant-based bev. with protein ≥ 1 RA/day | 185 (22.9%) | 173 (23.5%) | 160 (22.2%) | 134 (20.1%) |
| Unsaturated oils and fats ≥ 1 RA/day | 314 (38.8%) | 292 (39.6%) | 258 (35.8%) | 223 (33.4%) |
| **Adhering to Canada's Food Guide + additional protein foods – Females** | | | | |
| Protein foods, plant-based ≥ 1.3 RA/day | 61 (7.5%) | 51 (6.9%) | 58 (8.1%) | 55 (8.2%) |
| Protein foods, animal-based ≥ 3 RA/day | 169 (20.9%) | 138 (18.7%) | 119 (16.5%) | 99 (14.8%) |
| Milk and plant-based bev. with protein ≥ 1.5 RA/day | 74 (9.1%) | 71 (9.6%) | 72 (10.0%) | 68 (10.2%) |
| **Physical activity only – Females** | | | | |
| Physical activity ≥ 30 minutes/day | 203 (25.1%) | 146 (20.7%) | 143 (20.9%) | 125 (21.2%) |
| *^1^*Data in this table exclude values carried forward from previous follow-ups. Values are n (percentage). The proportions are based on mean data from all 24-h dietary recalls completed at each follow-up (up to three per follow-up). Since data were not corrected for random measurement error, the proportions do not reflect accurate group-level estimates of adherence to recommendations over a long period. Rather, the proportions should be interpreted as an assessment of the positivity assumption for the data used in the g formula algorithm. See Methods, Supplemental Methods - Sensitivity analysis to assess the impact of measurement error and the protocol article (Brassard et al. 2025) for details. For dietary intakes, 0, 139, 188 and 286 participants had missing values at baseline, follow-up 1, follow-up 2 and end of follow-up, respectively. For physical activity, 0, 177, 242 and 408 participants had missing values at baseline, follow-up 1, follow-up 2 and end of follow-up, respectively. RA, reference amounts. | | | | |

#### Simulated intervention data

***Table S8***: Pooled simulated dietary intakes ('intervened on') among all follow-ups in eligible NuAge participants*^1^*

|  | **Natural course (no intervention)** | | **CFG adherence with additional protein foods and physical activity** | | | |
| --- | --- | --- | --- | --- | --- | --- |
| **Characteristic** | **Females** N = 3,236 | **Males** N = 3,008 | **Females, excused** N = 285*^2^* | **Females, intervened** N = 2,951 | **Males, excused** N = 272*^2^* | **Males, intervened** N = 2,736 |
| Vegetables and fruits | 4.4 (2.0); [0.0-13.8] | 4.4 (2.1); [0.0-14.1] | 4.8 (1.9); [0.0-10.5] | 5.7 (1.1); [5.0-13.8] | 5.2 (1.8); [0.9-10.1] | 6.4 (0.9); [6.0-14.1] |
| Whole-grain foods | 0.8 (0.6); [0.0-3.5] | 1.0 (0.7); [0.0-4.4] | 1.3 (0.7); [0.0-3.6] | 1.7 (0.3); [1.5-3.5] | 1.4 (0.6); [0.0-3.5] | 1.7 (0.4); [1.5-4.4] |
| Protein foods, plant-based | 0.5 (0.7); [0.0-3.4] | 0.6 (0.9); [0.0-3.4] | 1.2 (1.1); [0.0-3.4] | 1.6 (0.6); [1.3-3.4] | 1.4 (1.2); [0.0-3.4] | 1.8 (0.7); [1.5-3.4] |
| Protein foods, animal-based | 2.2 (1.1); [0.0-6.9] | 2.4 (1.2); [0.0-6.9] | 2.3 (1.1); [0.0-4.9] | 3.2 (0.4); [3.0-6.9] | 2.5 (1.2); [0.0-6.2] | 3.6 (0.4); [3.5-6.9] |
| Milk and plant-based beverage with protein | 0.7 (0.6); [0.0-3.4] | 0.7 (0.6); [0.0-3.4] | 1.2 (0.5); [0.0-3.4] | 1.6 (0.3); [1.5-3.4] | 1.3 (0.5); [0.1-2.4] | 1.6 (0.3); [1.5-3.4] |
| Unsaturated oils and fats | 1.0 (1.1); [0.0-4.7] | 1.0 (1.1); [0.0-4.7] | 1.2 (1.2); [0.0-4.7] | 1.4 (0.9); [1.0-4.7] | 1.0 (1.0); [0.0-4.7] | 1.4 (0.8); [1.0-4.7] |
| Light or higher intensity physical activity, minutes/day | 18 (28); [0-303] | 28 (47); [0-437] | 28 (27); [0-164] | 42 (27); [30-331] | 41 (46); [0-269] | 52 (43); [30-437] |
| *^1^*Values are mean (SD); [Minimum-Maximum]. Data in this table include values carried forward from previous follow-ups. All dietary intakes are expressed in reference amounts per day. Among 'intervened' participants, values were modified to ensure that their intakes were consistent with the corresponding thresholds. See Table S1 for complete intervention details. CFG, Canada's Food Guide. | | | | | | |
| *^2^*Excused participants (incident cancer diagnosis, hospitalization, bed rest) were allowed to 'discontinue' the hypothetical intervention. That is, no hypothetical changes were made to their dietary intakes or amount of physical activity. However, the participants were not excluded from the analysis to avoid confounding by disease burden. See *Methods - Statistical Analysis for details*. | | | | | | |

#### Observed value of outcomes

**Table S9.** Observed values of outcomes and missing data across follow-ups in eligible NuAge participants^1^

| **Characteristic** | **Baseline** N = 1,561 | **Follow-up 1** N = 1,488 | **Follow-up 2** N = 1,390 | **End of follow-up** N = 1,285 |
| --- | --- | --- | --- | --- |
| Handgrip strength, kPa | 68.7 (18.8) | 66.2 (18.5) | 64.6 (18.5) | 62.0 (18.3) |
| (Missing) | 1 | 105 | 51 | 72 |
| Elbow flexor strength, kg | 20.0 (7.6) | 19.5 (7.5) | 18.4 (7.1) | 17.9 (7.1) |
| (Missing) | 3 | 124 | 102 | 110 |
| Quadriceps strength, kg | 26.9 (10.3) | 26.0 (9.9) | 24.8 (9.6) | 24.5 (9.6) |
| (Missing) | 10 | 99 | 115 | 132 |
| Timed Up & Go, seconds | 10.53 (2.29) | 10.87 (2.26) | 10.93 (2.77) | 11.01 (2.62) |
| (Missing) | 3 | 123 | 77 | 93 |
| Normal walking, best time, m/s | 1.17 (0.22) | 1.16 (0.22) | 1.16 (0.23) | 1.17 (0.23) |
| (Missing) | 3 | 118 | 75 | 92 |
| Fast walking, m/s | 1.50 (0.29) | 1.48 (0.30) | 1.47 (0.31) | 1.48 (0.31) |
| (Missing) | 5 | 119 | 77 | 92 |
| Waist circumference, cm | 95.4 (12.9) | 95.3 (12.9) | 95.9 (12.9) | 96.7 (13.1) |
| (Missing) | 3 | 106 | 50 | 71 |
| Systolic blood pressure, mmHg | 133 (17) | 130 (16) | 128 (16) | 126 (16) |
| (Missing) | 74 | 107 | 51 | 65 |
| Diastolic blood pressure, mmHg | 69 (9) | 72 (11) | 71 (9) | 70 (10) |
| (Missing) | 74 | 107 | 53 | 66 |
| Glomerular filtration rate, mL/min/1.73m2 | 74 (14) | 72 (14) | 71 (14) | 70 (15) |
| (Missing) | 212 | 246 | 179 | 216 |
| Glucose, mmol/L | 5.56 (1.28) | 5.53 (1.10) | 5.48 (1.06) | 5.31 (1.23) |
| (Missing) | 31 | 196 | 206 | 219 |
| 3MS total score, points | 94.0 (4.3) | 92.6 (5.8) | 92.8 (6.2) | 92.5 (5.9) |
| (Missing) | 6 | 98 | 38 | 95 |
| *^1^*Values are mean (SD). Of note, in main analyses (g formula algorithm), only the end of follow-up value was used as outcome. | | | | |

#### Main results, by sex

##### Muscle strength

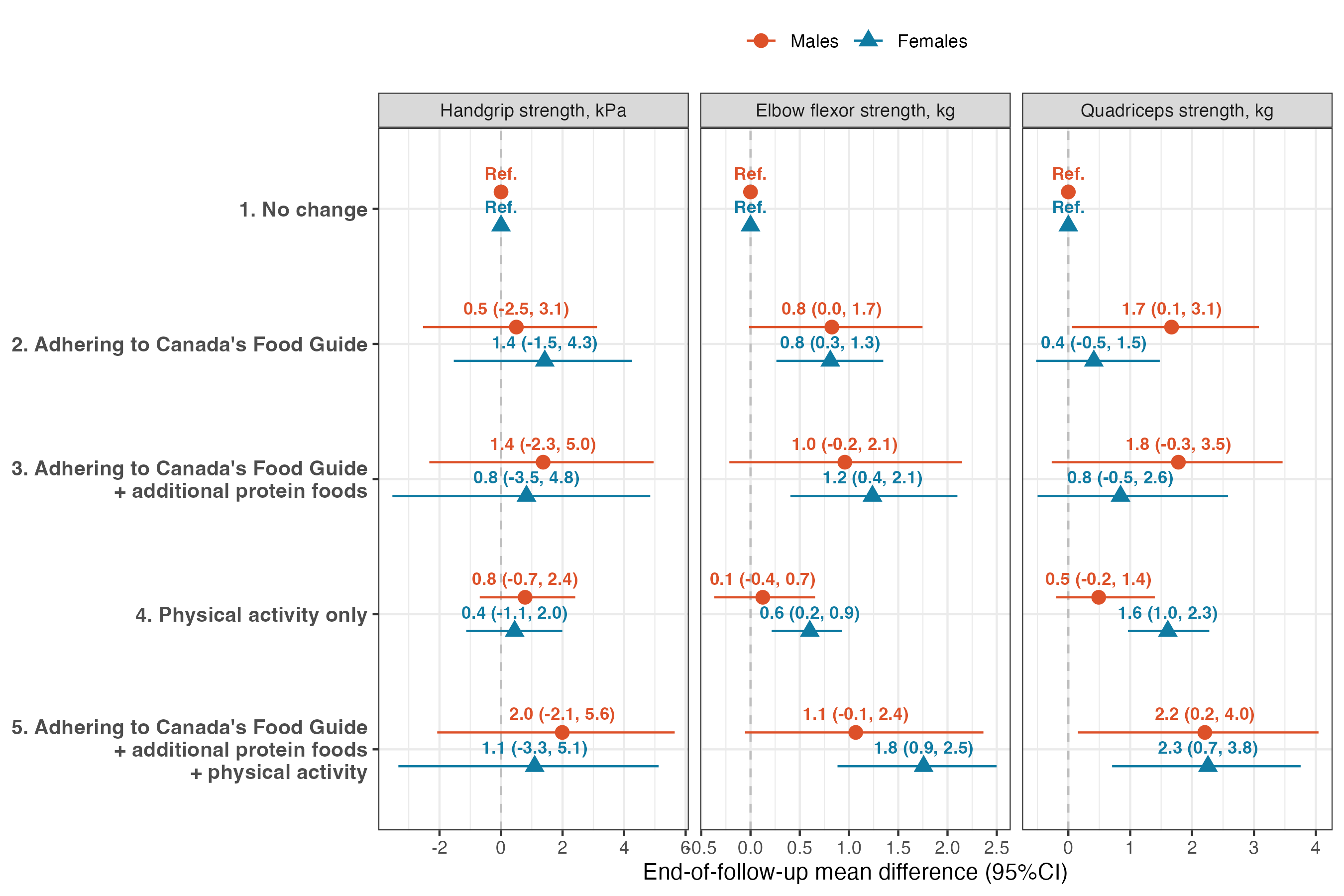

**Figure S2:** Mean difference in muscle strength outcomes after the 3-year hypothetical interventions, compared with no intervention, in eligible NuAge participants, stratified by sex. Effect estimates were obtained using the g-formula algorithm (see Methods). Physical activity corresponds to that of light intensity or higher. Models were adjusted for the covariates age, hormone replacement therapy, region, education, height, cancer prior to baseline, water and other healthy beverage intakes, total dietary intakes, living alone, measured body weight, number of medications, vitamin, mineral or natural health product consumption, alcohol consumption in the last month, current or former smoker, self-reported diabetes, self-reported high blood pressure, self-reported heart problems, as well as incident hospitalization, bed rest or cancer

##### Muscle function

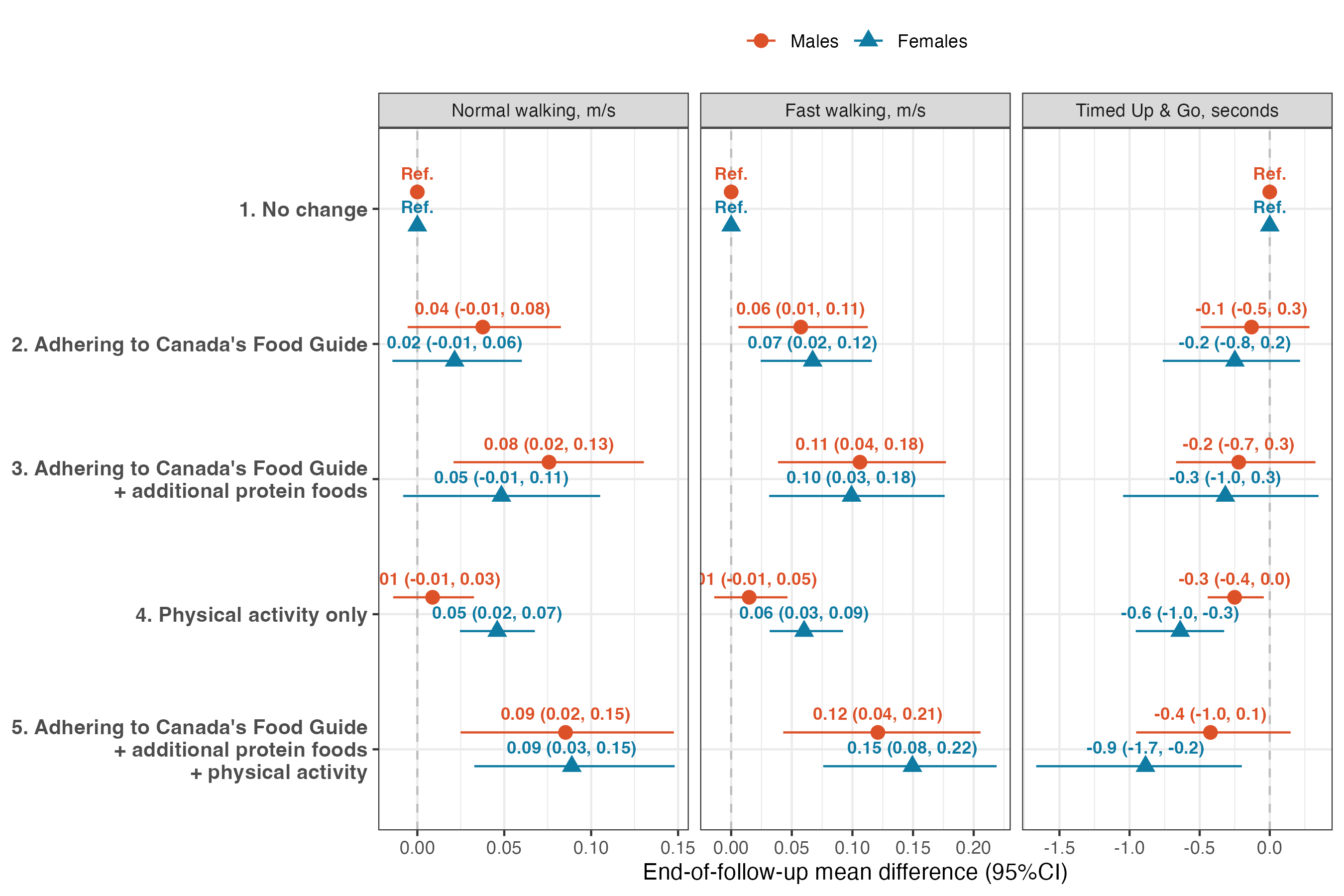

**Figure S3:** Mean difference in muscle function outcomes after the 3-year hypothetical interventions, compared with no intervention, in eligible NuAge participants, stratified by sex. Effect estimates were obtained using the g-formula algorithm (see Methods). Physical activity corresponds to that of light intensity or higher. Models were adjusted for the covariates age, hormone replacement therapy, region, education, height, cancer prior to baseline, water and other healthy beverage intakes, total dietary intakes, living alone, measured body weight, number of medications, vitamin, mineral or natural health product consumption, alcohol consumption in the last month, current or former smoker, self-reported diabetes, self-reported high blood pressure, self-reported heart problems, as well as incident hospitalization, bed rest or cancer

##### Cardiometabolic and Cognitive Health

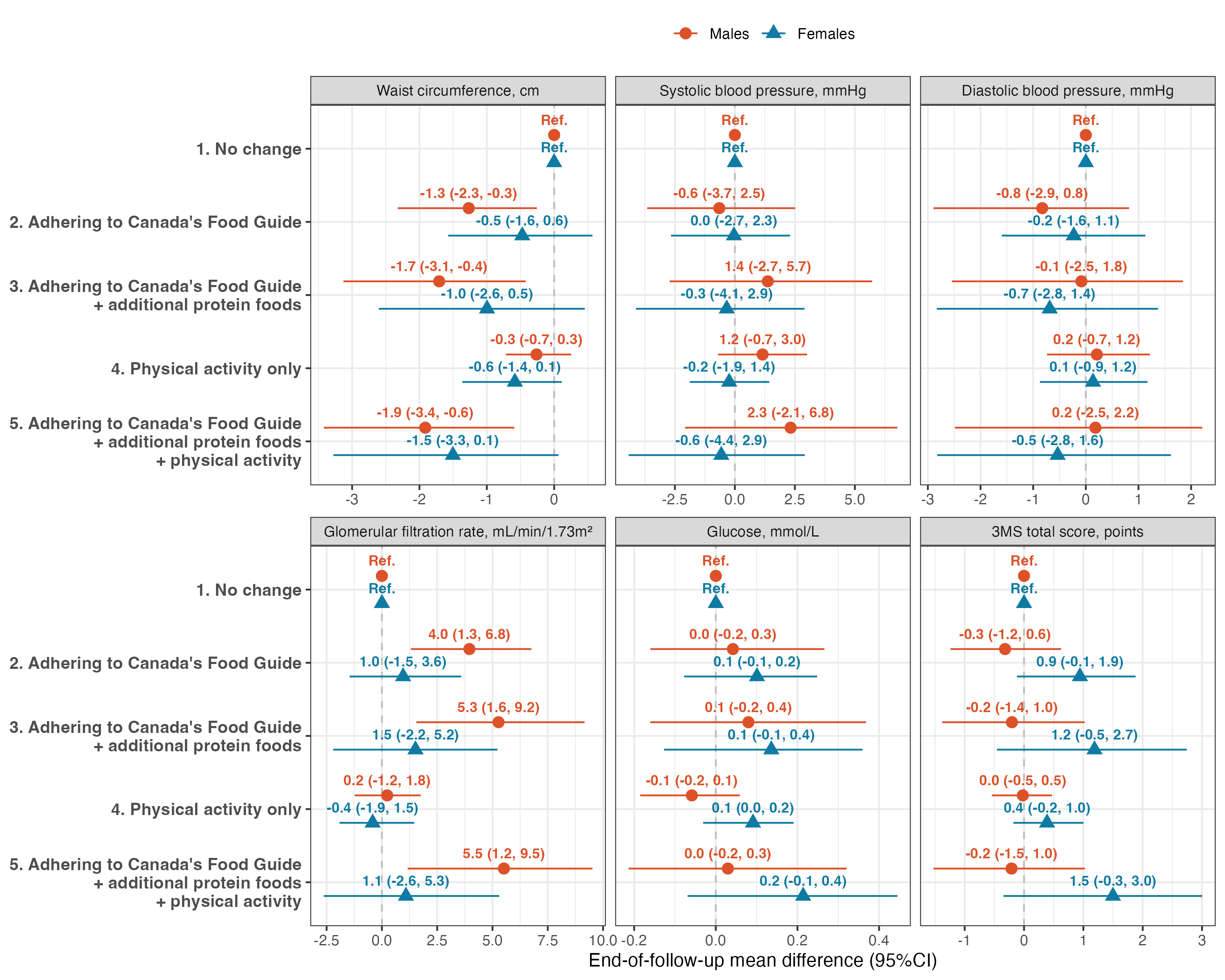

**Figure S4:** Mean difference in cardiometabolic health and cognition outcomes after the 3-year hypothetical interventions, compared with no intervention, in eligible NuAge participants, stratified by sex. Effect estimates were obtained using the g-formula algorithm (see Methods). Physical activity corresponds to that of light intensity or higher. Models were adjusted for the covariates age, hormone replacement therapy, region, education, height, cancer prior to baseline, water and other healthy beverage intakes, total dietary intakes, living alone, measured body weight, number of medications, vitamin, mineral or natural health product consumption, alcohol consumption in the last month, current or former smoker, self-reported diabetes, self-reported high blood pressure, self-reported heart problems, as well as incident hospitalization, bed rest or cancer

#### Sensitivity analyses

##### Assessment of unmeasured confounding

Table S10 presents the estimated effects of the hypothetical intervention on end of follow-up albumin concentrations. Albumin concentrations should not be strongly influenced by dietary intakes or physical activity, i.e., the hypothetical intervention under study. However, the presence of strong unmeasured or residual confounding by disease burden could create a spurious relationship between the hypothetical diet and physical activity intervention and albumin concentrations. Of note, the absence of relationships with albumin does not indicate that there is no unmeasured or residual confounding.

**Table S10.** Estimated effects of the hypothetical intervention on end of follow-up albumin concentrations (g/L) in eligible NuAge participants^1^

| **Hypothetical intervention** | **Mean end of follow-up treatment value** | **Difference of mean treatment value (95%CI)** |
| --- | --- | --- |
| **All participants (n=1561)** | | |
| No intervention | 40.6 | Reference |
| Adherence to CFG | 40.8 | 0.2 (-0.1, 0.6) |
| Adherence to CFG with additional protein foods | 40.8 | 0.2 (-0.3, 0.7) |
| Physical activity only | 40.5 | -0.1 (-0.3, 0.1) |
| Adherence to CFG with additional protein foods and physical activity | 40.7 | 0.1 (-0.4, 0.6) |
| **Males only (n=752)** | | |
| No intervention | 40.7 | Ref. |
| Adherence to CFG | 40.8 | 0.1 (-0.5, 0.6) |
| Adherence to CFG with additional protein foods | 40.9 | 0.2 (-0.6, 0.9) |
| Physical activity only | 40.4 | -0.2 (-0.5, 0.2) |
| Adherence to CFG with additional protein foods and physical activity | 40.7 | 0.1 (-0.8, 0.8) |
| **Females only (n=809)** | | |
| No intervention | 40.4 | Ref. |
| Adherence to CFG | 40.8 | 0.4 (-0.1, 1.0) |
| Adherence to CFG with additional protein foods | 40.7 | 0.3 (-0.5, 1.3) |
| Physical activity only | 40.6 | 0.2 (-0.1, 0.5) |
| Adherence to CFG with additional protein foods and physical activity | 40.9 | 0.5 (-0.4, 1.4) |
| *^1^*Values were estimated using the parametric g-formula (see Methods). 'No intervention' corresponds to the estimated albumin concentration when dietary intakes and the amount of physical activity are not modified. Variance was estimated using 500 bootstrap resamples. Sex-specific estimates were obtained by stratification. Models were adjusted for age, sex (all participants model only), hormone replacement therapy, region, education, height, cancer prior to baseline, water and other healthy beverage intakes, total dietary intakes, living alone, measured body weight, number of medications, vitamin, mineral or natural health product consumption, alcohol consumption in the last month, current or former smoker, self-reported diabetes, self-reported high blood pressure, self-reported heart problems, as well as incident hospitalization, bed rest or cancer. CFG, Canada's Food Guide. | | |

##### Assessment of the impact of measurement error

Dietary intakes measured with 24-h dietary recalls are less prone to systematic error than other instruments (e.g., food-frequency questionnaire), but are affected by within-individual random errors. In the present study, the mean dietary intakes based on all 24-h dietary recalls completed by each participant (up to three follow-up) were calculated to mitigate random errors, at least to some extent. However, residual random errors are expected and may bias effect estimate. The National Cancer Institute Markov Chain Monte Carlo multivariate method was used to obtain measurement error-corrected effect estimates for comparison purpose.

**Table S11.** Observational analog of the intention-to-treat effect estimates of within-person mean dietary intakes compared with estimated usual intakes^1^

|  | **Effect estimates (95%CI)** | | **Bias estimate** | |
| --- | --- | --- | --- | --- |
| **Outcomes** | Estimated usual intakes*^2^* | Within-person mean*^3^* | Difference vs. usual | Ratio vs. usual*^4^* |
| **Muscle strength** | | | | |
| Handgrip strength, kPa | 2.5 (-4.6, 9.8) | 2.3 (-0.3, 4.7) | -0.22 | 0.91 |
| Elbow flexor strength, kg | 0.0 (-1.4, 1.6) | 0.3 (-0.3, 0.9) | 0.34 | NA |
| Quadriceps strength, kg | 3.0 (0.4, 5.8) | 1.6 (0.6, 2.7) | -1.39 | 0.54 |
| **Muscle function** | | | | |
| Timed Up & Go, seconds | -1.3 (-2.5, -0.1) | -0.5 (-1.0, -0.2) | 0.79 | 0.41 |
| Normal walking, m/s | 0.06 (-0.03, 0.16) | 0.04 (0.01, 0.07) | -0.02 | 0.72 |
| Fast walking, m/s | 0.07 (-0.05, 0.21) | 0.05 (0.01, 0.09) | -0.02 | 0.74 |
| **Cardiometabolic and cognitive health** | | | | |
| Waist circumference, cm | -1.3 (-3.8, 0.8) | -0.8 (-1.7, -0.1) | 0.44 | 0.66 |
| Systolic blood pressure, mmHg | -4.6 (-11.0, 0.8) | -2.6 (-5.1, -0.6) | 1.99 | 0.57 |
| Diastolic blood pressure, mmHg | -1.0 (-5.6, 3.0) | -0.8 (-2.5, 0.6) | 0.19 | 0.81 |
| Glomerular filtration rate, mL/min/1.73m² | 1.0 (-4.3, 8.1) | 1.4 (-0.7, 4.0) | 0.45 | 1.46 |
| Glucose, mmol/L | 0.0 (-0.4, 0.4) | 0.0 (-0.2, 0.1) | -0.02 | NA |
| 3MS total score, points | 0.9 (-1.3, 3.4) | 0.2 (-0.7, 1.1) | -0.65 | 0.28 |
| **Negative control** | | | | |
| Albumin, g/L | 0.0 (-1.2, 1.3) | 0.1 (-0.4, 0.6) | 0.05 | NA |
| *^1^*Estimates reflect end of follow up outcome difference after a hypothetical intervention 'assigned' at baseline only, compared with no intervention. The hypothetical intervention corresponds to adherence to Canada's Food Guide. Only the baseline value of time-varying dietary intakes, physical activity and covariates was considered. Models were adjusted for age, sex, hormone replacement therapy, region, education, height, cancer prior to baseline, water and other healthy beverage intakes, total dietary intakes, living alone, measured body weight, number of medications, vitamin, mineral or natural health product consumption, alcohol consumption in the last month, current or former smoker, self-reported diabetes, self-reported high blood pressure, as well as self-reported heart problems. Pre-baseline dietary intakes were also included as covariates. NA, not applicable (the reference estimate is null). | | | | |
| *^2^*Usual dietary intakes were estimated using the National Cancer Institute Markov Chain Monte Carlo multivariate method. These effect estimates are corrected for (within-person) random measurement errors using regression calibration. Variance was estimated using 300 bootstrap resamples. | | | | |
| *^3^*Within-person mean corresponds to the mean dietary intakes among all 24-h dietary recalls completed by each participant (same as in the main analysis). Residual random measurement errors associated with 24-h dietary recalls may introduce bias to effect estimates. Variance was estimated using 300 bootstrap resamples. | | | | |
| *^4^*Interpretation: a ratio greater than 1 indicates that effect estimates based on within-person mean were overestimated, and vice-versa. A negative sign indicates that effect estimates were in the opposite direction. For example, the effect of the hypothetical intervention for Timed Up & Go time was 0.41-fold smaller using within-person mean intakes compared with usual intakes, while in the same direction. | | | | |

##### Assessment of model misspecification

Figure S5 presents the comparison between observed (“IP weighted estimates”; natural course accounting for loss to follow-up) and predicted values (“parametric g formula estimates”) of time-varying variables.

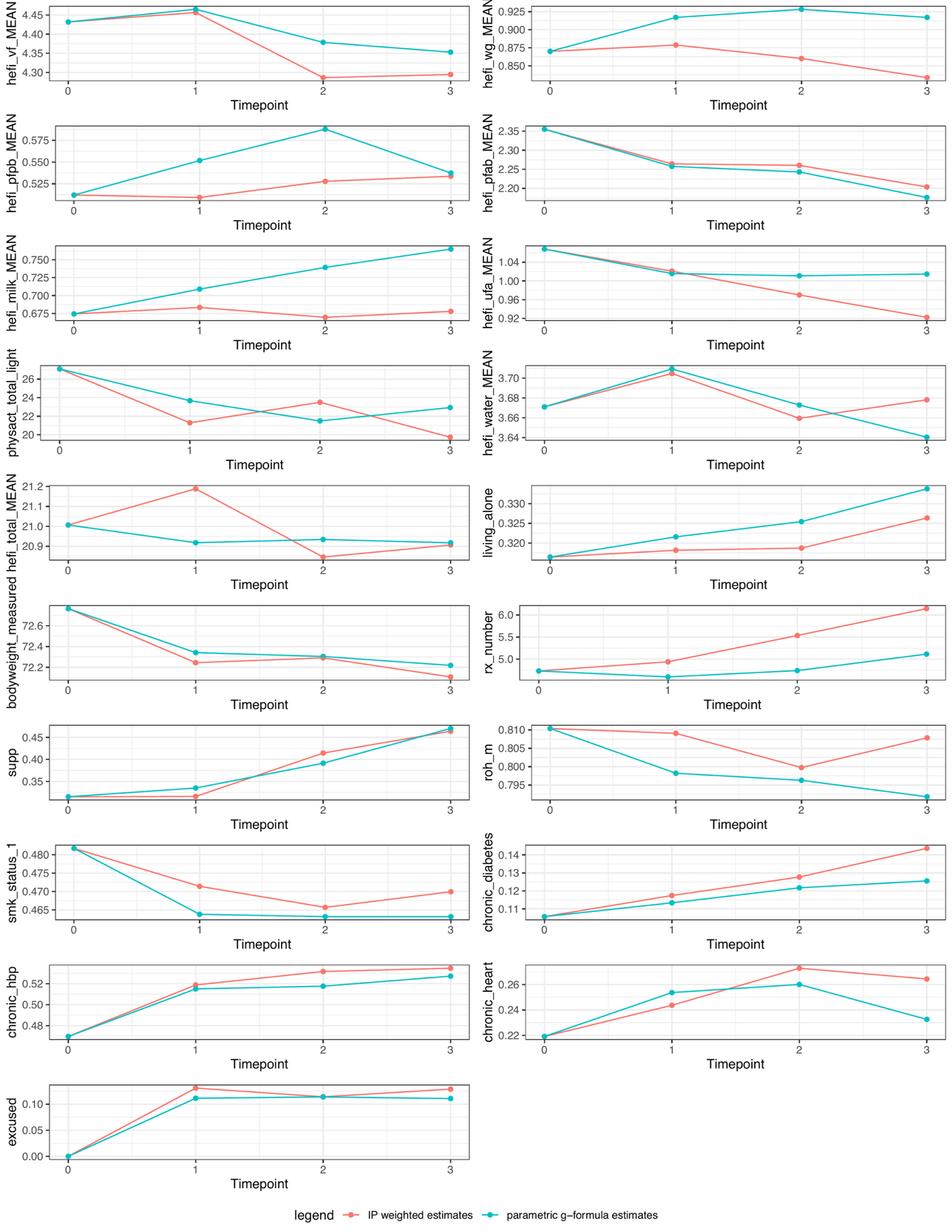

**Figure S4:** Assessment of time-varying models misspecification in the g formula algorithm. Divergence between the parametric g−formula estimates and the inverse-probability weighted estimates suggest that the functional form of covariates and/or modelling of outcomes may have been incorrect. Variables definition: bodyweight_measured, measured body weight; chronic_diabetes, self-reported diabetes; chronic_hbp, self-reported high blood pressure; chronic_heart, self-reported heart problems; excused, incident hospitalization, bed rest or cancer; hefi_milk_MEAN, unsweetened milk; hefi_pfab_MEAN, animal-based protein foods; hefi_pfpb_MEAN, plant-based protein foods; hefi_total_MEAN, total dietary intakes; hefi_ufa_MEAN, unsaturated fats and oils; hefi_vf_MEAN, vegetables and fruits; hefi_water_MEAN, water and other healthy beverages; hefi_wg_MEAN, whole-grain foods; IP, inverse probability; living_alone, living alone; physact_total_light, total light or higher intensity activities; roh_m, alcohol consumption in the last month; rx_number, number of medications; smk_status_1, current or former smoker; supp, vitamin, mineral or natural health product consumption.

#### References

Brassard, D., L. A. Elvidge Munene, S. St-Pierre, A. Gonzalez, P. M. Guenther, M. Jessri, J. Vena, et al. 2022. “Evaluation of the Healthy Eating Food Index (HEFI)-2019 Measuring Adherence to Canada’s Food Guide 2019 Recommendations on Healthy Food Choices.” Journal Article. *Appl Physiol Nutr Metab* 47 (5): 582–94. <https://doi.org/10.1139/apnm-2021-0416>.

Brassard, D., L. A. Elvidge Munene, S. St-Pierre, P. M. Guenther, S. I. Kirkpatrick, J. Slater, S. Lemieux, et al. 2022. “Development of the Healthy Eating Food Index (HEFI)-2019 Measuring Adherence to Canada’s Food Guide 2019 Recommendations on Healthy Food Choices.” Journal Article. *Appl Physiol Nutr Metab* 47 (5): 595–610. <https://doi.org/10.1139/apnm-2021-0415>.

Brassard, D., N. Presse, and S. Chevalier. 2025. “Estimating the Effect of Adhering to the Recommendations of the 2019 Canada’s Food Guide on Health Outcomes in Older Adults: Protocol for a Target Trial Emulation.” Journal Article. *JMIR Res Protoc*, 14:e65182. <https://doi.org/10.2196/65182>.

Desrosiers, J., F. Prince, A. Rochette, and M. Raîche. 1998. “Reliability of Lower Extremity Strength Measurements Using the Belt-Resisted Method.” Journal Article. *Journal of Aging and Physical Activity* 6 (4): 317–26. <https://doi.org/10.1123/japa.6.4.317>.

Health Canada. 2022. “Table of Reference Amounts for Food.” Web Page. Ottawa: Health Canada. <https://www.canada.ca/en/health-canada/services/technical-documents-labelling-requirements/table-reference-amounts-food.html>.

Hernán, M. A. 2021. “Causal Analyses of Existing Databases: No Power Calculations Required.” Journal Article. *J Clin Epidemiol*. <https://doi.org/10.1016/j.jclinepi.2021.08.028>.

Hernán, M. A., and J. M. Robins. 2020. *Causal Inference: What If*. Book. CRC Boca Raton: Chapman & Hall/CRC. [https://www.hsph.harvard.edu/miguel-hernan/causal-inference-book/](%20https://www.hsph.harvard.edu/miguel-hernan/causal-inference-book/).

Keogh, R. H., P. A. Shaw, P. Gustafson, R. J. Carroll, V. Deffner, K. W. Dodd, H. Kuchenhoff, et al. 2020. “STRATOS Guidance Document on Measurement Error and Misclassification of Variables in Observational Epidemiology: Part 1-Basic Theory and Simple Methods of Adjustment.” Journal Article. *Stat Med* 39 (16): 2197–2231. <https://doi.org/10.1002/sim.8532>.

Kirkpatrick, S. I., P. M. Guenther, A. F. Subar, S. M. Krebs-Smith, K. A. Herrick, L. S. Freedman, and K. W. Dodd. 2022. “Using Short-Term Dietary Intake Data to Address Research Questions Related to Usual Dietary Intake Among Populations and Subpopulations: Assumptions, Statistical Techniques, and Considerations.” Journal Article. *J Acad Nutr Diet* 122 (7): 1246–62. <https://doi.org/10.1016/j.jand.2022.03.010>.

McGrath, Sean, Victoria Lin, Zilu Zhang, Lucia C. Petito, Roger W. Logan, Miguel A. Hernán, and Jessica G. Young. 2020. “gfoRmula: An r Package for Estimating the Effects of Sustained Treatment Strategies via the Parametric g-Formula.” Journal Article. Patterns 1 (3). <https://doi.org/10.1016/j.patter.2020.100008>.

Payette, H., N. Hanusaik, V. Boutier, J. A. Morais, and K. Gray-Donald. 1998. “Muscle Strength and Functional Mobility in Relation to Lean Body Mass in Free-Living Frail Elderly Women.” Journal Article. *Eur J Clin Nutr* 52 (1): 45–53. <https://doi.org/10.1038/sj.ejcn.1600513>.

Podsiadlo, D., and S. Richardson. 1991. “The Timed "up & Go": A Test of Basic Functional Mobility for Frail Elderly Persons.” Journal Article. *J Am Geriatr Soc* 39 (2): 142–48. <https://doi.org/10.1111/j.1532-5415.1991.tb01616.x>.

Teng, E. L., and H. C. Chui. 1987. “The Modified Mini-Mental State (3MS) Examination.” Journal Article. *J Clin Psychiatry* 48 (8): 314–18. <https://www.ncbi.nlm.nih.gov/pubmed/3611032>.

Thompson, F. E., S. I. Kirkpatrick, A. F. Subar, J. Reedy, T. E. Schap, M. M. Wilson, and S. M. Krebs-Smith. 2015. “The National Cancer Institute’s Dietary Assessment Primer: A Resource for Diet Research.” Journal Article. *J Acad Nutr Diet* 115 (12): 1986–95. <https://doi.org/10.1016/j.jand.2015.08.016>.

Zhang, S., D. Midthune, P. M. Guenther, S. M. Krebs-Smith, V. Kipnis, K. W. Dodd, D. W. Buckman, J. A. Tooze, L. Freedman, and R. J. Carroll. 2011. “A New Multivariate Measurement Error Model with Zero-Inflated Dietary Data, and Its Application to Dietary Assessment.” Journal Article. *Ann Appl Stat* 5 (2B): 1456–87. <https://doi.org/10.1214/10-AOAS446>.
